# The effect of Behavioral Factors and Intervention Strategies on Pathogen Transmission: Insights from a Two-Week Epidemic Game at Wenzhou-Kean University in China

**DOI:** 10.1101/2024.12.14.24318955

**Authors:** Salihu S. Musa, Winnie Mkandawire, Trusting Inekwe, Yinan Dong, Andonaq Grozdani, Hung Hong, Mansi Khandpekar, Sarah A. Nowak, Jean-Gabriel Young, Aloysius Wong, Dale King, Andrés Colubri

## Abstract

**Background:** Effective control of infectious diseases relies heavily on understanding transmission dynamics and implementing interventions that reduce the spread. Non-pharmaceutical interventions (NPIs), such as mask-wearing, social distancing, and quarantining, are vital tools in managing outbreaks where vaccines or treatments are limited. However, the success of NPIs is influenced by human behavior, including compliance with guidelines, and attitudes such as beliefs about the effectiveness of interventions. In this study, we applied an innovative proximity-based experimentation platform to generate empirical data on behaviors and attitudes and their effect on disease transmission. Our platform uses a smartphone application that enables the spread of a digital pathogen among participants via Bluetooth during open-world “experimental epidemic games”. This creates an environment for epidemiology field experimentation where researchers can control transmission mechanics and collect full ground-truth datasets.

**Methods:** Our study employed the “epidemic” app to investigate the impact of risk perception and compliance to NPIs on pathogen transmission. Involving nearly 1,000 participants in a two-weeks long epidemic game at Wenzhou-Kean University (WKU) in China, the app generated a multimodal dataset, which allowed us to develop and parameterize Susceptible-Exposed-Infected-Recovered (SEIR) models. We quantified the extent by which behavioral factors, such as risk perception and compliance with quarantine, and strength of intervention strategies influence disease transmission. The model incorporates time-varying transmission rates that reflect changes in attitudes and behavior, and we calibrated it using the empirical data from the epidemic game to provide critical insights into how variations in NPI compliance levels affect outbreak control.

**Findings:** The findings reveal that adherence to NPIs alone, which is influenced by changes in behavior and attitudes, may not result in the expected reduction in transmission, illustrating the complex interplay between behavioral factors and epidemic control. Moreover, the model further shows that changes in risk perception coupled with NPI adherence could significantly reduce infection levels as well as susceptibility.

**Interpretation:** Our study highlights the usefulness of experimental epidemic games to generate realistic datasets, and the importance of integrating behavioral dynamics into epidemiological models to enhance the accuracy of predictions and the effectiveness of public health interventions during infectious disease outbreaks.

**Research in Context:** *Evidence before this study:* We conducted a comprehensive review of the existing literature to evaluate the current state of knowledge regarding empirically-informed infectious disease modeling, with a particular focus on the role of human behavior and non-pharmaceutical interventions (NPIs) in mitigating disease transmission. Our search spanned databases such as PubMed, MEDLINE, and Web of Science, targeting publications up to March 1, 2024, using keywords including “infectious disease modeling,” “simulation,” “experimental game,” “human behavior,” “non-pharmaceutical interventions,” and “epidemiology.” While a substantial body of research explores the influence of human behavior on disease dynamics, there is a notable gap in studies that integrate large-scale mobility and behavioral data collected with smartphone apps within open-world environments, such as a university campus. Most existing studies fail to incorporate the complexity of real-time human behavioral responses and NPIs, which are crucial for accurately modeling the dynamics of disease transmission in such contexts.

*Added value of this study:* This study is the first to use our proximity-based experimentation platform to conduct an epidemic game in a large-scale university setting while integrating human behavioral factors and NPIs into a mechanistic modeling framework. By employing a flexible, time-varying transmission rate model, our research highlights the impact of human behavior and NPIs on pathogen spread dynamics. This novel approach provides a more accurate and nuanced depiction of real-world transmission scenarios, as observed during the proximity-based experiment. Through the integration of empirical data from nearly 1,000 participants, combined with detailed model simulations and rigorous sensitivity analyses, we offer insights into how timely and coordinated interventions, alongside public compliance, can significantly influence the trajectory of an outbreak. This study underscores the necessity of adaptive strategies in outbreak management and presents a robust framework that can inform and enhance future public health planning and response efforts.

*Implications of all the available evidence:* Our findings underscore the pivotal role of experimental and computational approaches for generating realistic outbreak datasets and integrating behavioral dynamics and NPIs into epidemiological models. This results in significantly more accurate models that then can become valuable tools for public health planning. The study provides a solid foundation for refining models with additional complexities, such as age-based behaviors, and offers a framework for optimizing outbreak management and future pandemic preparedness.

## 1 Introduction

The advent of new digital technologies has significantly transformed infectious disease research, including both epidemiological data collection and modeling. In particular, mobile applications (apps) can leverage the sensing and communication capabilities of smartphones and wearable devices for generating large amounts of real-time data at multiple levels. During the COVID-19 pandemic, apps were used for participatory surveillance, population-level tracking, individual risk assessment, individual screening, digital contact tracing, and education [1]. Several years before the pandemic, we started working on a proximity-based app called Operation Outbreak (OO) that facilitates immersive engagement in “mock” outbreaks that take place in real-life settings, such as schools and conferences, enhancing our understanding of outbreak dynamics and response strategies thanks to its capacity to incorporate naturalistic human behavior and controlled transmission mechanics [2, 3]. The OO mobile application is freely accessible on Google and Apple app stores [4, 5], and it is supported by a scalable cloud backend and web-based administration and analytics tools. This app enables a digital infection that spreads through participants’ mobile phones via Bluetooth, triggering a “synthetic” outbreak that participants can respond realistically to by endowing the app with appropriate reward or incentive mechanisms to various participants’ actions. The early example of the “Corrupted Blood incident” virtual pandemic in the World of Warcraft massive multiplayer online role playing game (MMORPG) back in 2007 prompted epidemiologists [6] to consider MMORPGs and other virtual games as settings where individual human behavior in response to pathogen spread could be studied experimentally rather than via modeling assumptions, issues of external validity notwithstanding [7]. The OO app was initially motivated by infectious disease education and pandemic preparedness, as the original version of the app was designed to support an immersive educational experience in middle and high schools centered around public health, outbreak response, and societal roles during health emergencies [8]. While this continues to be one of the main thrusts of the project [12], here we used a customized version of the OO app and backend to more specifically target epidemiology experimentation and research [18], by generating valuable data that informs epidemiological models and providing insights into pathogen transmission dynamics [19, 20, 21, 22].

In this study, we conducted a proximity-based “experimental epidemic game” at the campus of Wenzhou-Kean University (WKU), an international Chinese-American institution of highereducation established by Kean University of New Jersey in the Zhejiang province of eastern China [9], which we will call the WKU game (or experiment) for short for the remainder of the paper. The aims of the WKU game were to generate a comprehensive outbreak dataset, explore spread of the simulated pathogen, and assess behavioral responses within the naturalistic environment of a college campus given a ground-truth transmission model in the app. Planning began in early fall 2023, with strategic recruitment of WKU students as organizers and facilitators. Our primary objectives were to engage at least 1,000 students over a two-week observational period and to integrate features representing individual behavioral actions within the experimentation framework. Drawing on methodologies from past (proximity-based) epidemic games at institutions like Colorado Mesa University and Brigham-Young University [3], we reached high participation rates (nearly a quarter of the enrolled students) through a comprehensive campus information campaign and incentivization strategies. In preparation for this experiment, we advanced the underlying technology in the app, including the incorporation of the open-source Herald proximity library [10] and migration to the Flutter framework [11], enhancing the app’s compatibility, adaptability and scalability [3].

Epidemiological modeling, when integrated with human behavior, serves as a foundational tool for evaluating the impact of non-pharmaceutical interventions (NPIs) on disease dynamics [3, 19, 21, 24]. The WKU proximity-based epidemic game combined open-world data with an epidemiological framework to study pathogen spread within a largely closed population. This interdisciplinary approach aims to contribute valuable insights into epidemiological modeling and public health preparedness, focusing on behavioral responses and the effectiveness of interventions in containing and mitigating disease outbreaks [3, 20, 23, 25]. Researchers have used online behavioral experiments to try to measure and model the effect of individuals’ preferences and decisions on adoption of protective measures and disease dynamics [26, 27, 28]; however, such experiments often reduce complex human behavior to simple abstract decisions that may not resonate with participants’ real-world concerns or priorities and may fail to reflect the complex social contexts and disease exposure patterns that influence behavior in realistic environments. Our approach has the advantage of providing an open-world experimentation framework with high degree of mechanistic realism for the transmission processes and a naturalistic environment able to capture inter-individual variation in real-world social settings [18]. The idea of conducting real-world “simulations” or experiments to study disease transmission has been explored before by projects like FluPhone [13] in 2010, and more recently, SafeBlues [14]. Both used Bluetooth sensing in mobile phones to spread virtual pathogens through a network of participants, but thanks to advances in mobile technology over the last decade, we were able to construct a new platform with a significantly greater potential for customization and scale than any earlier projects we are aware of.

Numerous studies have examined the impact of NPIs on pathogen transmission, providing critical insights into how measures such as social distancing, mask-wearing, and isolation can alter the course of an epidemic [29, 30]. For instance, SIRS models have demonstrated that the attributes of a pathogen, particularly its basic reproduction number (*R*_0_), significantly influence the long-term epidemiological outcomes of transmission reduction efforts [31, 32]. Pathogens with high *R*_0_ values may exhibit only temporary reductions in transmission rates, with minimal impact on the epidemic’s overall dynamics [41]. Additionally, other studies have highlighted the importance of seasonality and immunity in shaping medium-to long-term epidemic dynamics. These studies suggest that intermittent application of NPIs may lead to delayed but potentially larger secondary transmission peaks, underscoring the complexity of public health responses. The interaction between NPIs and pathogen characteristics necessitates ongoing evaluation and adjustment of intervention strategies to effectively address unexpected outcomes [32, 33, 34, 35].

Incorporating behavioral factors into epidemiological models significantly enhances our understanding of disease dynamics by elucidating the complex interplay between human behavior and pathogen transmission [30, 36]. Traditional models often overlook the variability in human behavior, treating responses to outbreaks as static or uniform. However, this approach does not fully account for the adaptive nature of individual and collective actions in response to evolving risk perceptions and public health interventions. Our study tries to address these challenges by integrating behavior as a dynamic variable, where individuals adjust their adherence to NPIs based on the perceived severity of the outbreak and their personal risk assessment (that is influenced by real-time factors such as disease prevalence, peer influence, and the effectiveness of public health communication) [30, 36]. Using the dataset generated from the WKU game, our model not only captures the immediate impact of NPIs such as social distancing and mask-wearing but also considers the feedback loops between behavioral adaptations and disease dynamics [37, 38]. This approach provides a more realistic and nuanced understanding of pathogen transmission, reflecting the complex interplay between human actions and epidemiological outcomes [36, 37]. While our model advances the field by incorporating these dynamic behavioral factors, we acknowledge the ongoing challenge of fully capturing the spectrum of human behavior, which underscores the need for continuous refinement and development in this area of research [39, 40].

Our modeling framework integrates these insights by utilizing data-driven approaches to simulate pathogen transmission and assess the impact of various interventions, such as quarantine, isolation, and face masks [24]. By leveraging mobile app technology, our platform allows for a detailed examination of how individual decisions—such as mask wearing, social distancing, or isolating when symptomatic—affect transmission dynamics within the controlled environment of the game, in turn embedded in the open-world setting of a college campus. The models were calibrated using data from the WKU game, enabling a precise analysis of transmission dynamics and the effectiveness of different interventions. This approach not only enhances our understanding of disease dynamics but also underscores the importance of timely interventions and compliance with preventive measures in mitigating outbreaks.

This study focuses on establishing a foundation for comprehensive models that can guide public health policies and strategies to prevent and reduce infectious disease outbreaks. The data generated from the WKU game was analyzed using a conceptual epidemiological modeling approach that integrates human attitudes, behaviors, and NPIs to disentangle and quantify the impact of these factors on pathogen spread. By fitting the model to the empirical data from our experimental game, we validated its accuracy and identified key parameters driving transmission dynamics. Sensitivity analyses and contour plots further highlight the importance of timely interventions, demonstrating how individual behavior and NPIs can significantly reduce infection rates. This research not only advances our theoretical understanding of socio-epidemiological interactions but also offers practical guidance for managing infectious diseases across diverse settings.

The paper is organized as follows: Section 2 describes the compartmental model we employed to mathematically represent disease transmission during the WKU game, which includes a variable transmission rate to capture adaptive behavioral factors and strength of NPIs. This section also calibrates the model with data from the two-week long WKU game. Section 3 provides an examination of the model that focuses on the occurrence and stability of disease-free and endemic equilibria, as well as the possibility of other complex dynamics. Results on the impact of behavior and NPIs are also presented in this section. Section 4 includes global uncertainty and parameter sensitivity analyses to evaluate the model’s robustness and identify critical parameters influencing transmission dynamics. Section 5 presents the results of numerical simulations and insights gained from global parameter sensitivity studies. Finally, Section 6 summarizes the study’s findings and discusses the implications for public health strategies and future research paths.

## 2 Methods

### 2.1 Experimental design

We designed the WKU experimental epidemic game to study human behavior and NPIs during an infectious disease outbreak. Experimental games constitute a novel approach that has been applied in fields such as public health and climate science and it is gaining recognition as a valuable source of behavioral data for research [16, 17, 15]. The app spreads the digital pathogen using an underlying “ground-truth” transmission model that assigns a probability of infection to each proximity contact between a susceptible and an infectious user (refer to the appendix for details) and informs users of their simulated health status (asymptomatic, mild and severely ill, recovered, and deceased) via an animated avatar and changes in the color of the app’s user interface (UI). A key difference with prior experimental games is the open-world, naturalistic setting of our experiment, which allows for a highly immersive experience and enables students to adopt behaviors during our experiment such as socially distancing to reduce their chances of infection, very closely mirroring real-life.

Furthermore, we considered how to gamify health-related decision-making in the app in such a way that students would need to make choices in the experiment on a regular basis, mirroring conflict between individual and group benefits, just like in a real outbreak. Based on these considerations, we implemented a point-based system in which students could collect points by deciding to ’quarantine / isolate’ or not at the beginning of each day and then use those points to purchase virtual masks and rapid diagnostic kits through a ’shop’ feature within the app. Of course, it would not have been reasonable to ask students to quarantine/isolate by physically restricting their movement, instead, they quarantined/isolated by selecting a button in the app, which made their avatars invisible to nearby participants (both quarantine and isolation used the same mechanics, the difference in using one term of the other refers to whether the student was in an asymptomatic state or not at the moment of their selection.) The goal of the point system was to provide a quick mechanism for students to get points without distracting them from their daily school activities, but still offering a sense of personal investment in the experiment, as their points informed a school-wide leaderboard that was used to award prizes to the top-scoring students once the experiment ended. See Fig. 1 for pictures of the students using the app and some screen captures depicting its main screen and quarantine/isolate feature.

**Figure 1:**
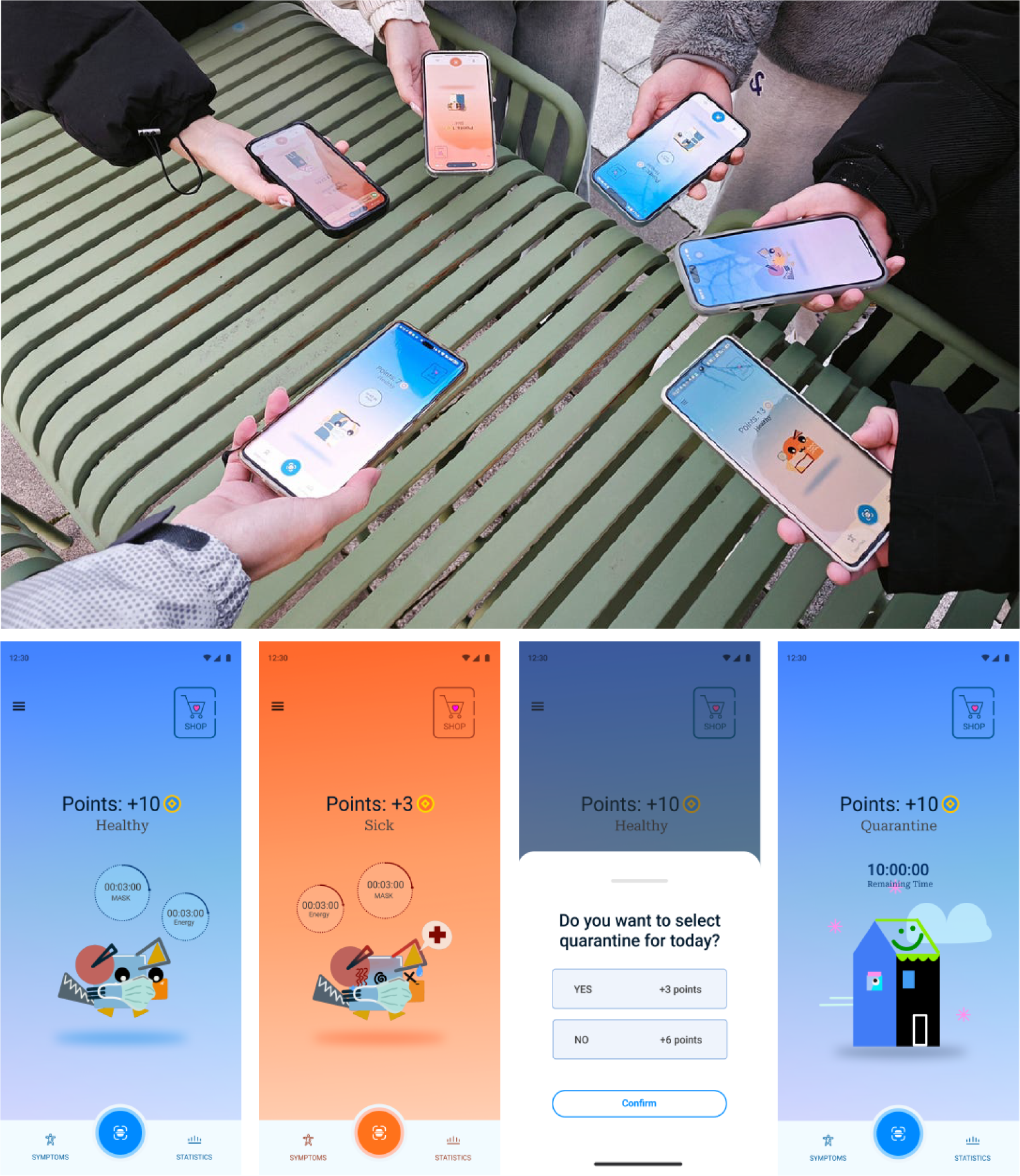
Students engaging with the smartphone app simulating pathogen transmission and disease progression during the epidemic game at Wenzhou-Kean University (WKU) (top). Screenshots of the app, showing (from left to right) the home page with healthy and sick avatars, a pop-up message prompting students to choose whether to quarantine/isolate for the day, and the home page interface while in quarantine/isolation (bottom). These visuals illustrate the interactive features of the app, which are integral to collecting behavioral and epidemiological data during the WKU game.

We also planned and implemented a pre-experiment stage to engage a substantial portion of the student body, employing a multifaceted strategy across various communication platforms (see Fig. 2). This strategy included the design of a dedicated informational website, the formation of WeChat groups for seamless communication, and the distribution of both digital and physical flyers to effectively disseminate information. A pivotal component of this strategy was the use of a registration form managed by the local organization team, allowing for the systematic collection of student details and facilitating insights into participation trends through regular updates. Integration with WeChat further enhanced communication channels, enabling efficient handling of inquiries and fostering active engagement among participants; this approach to data collection has proven effective [42, 43, 44]. Moreover, incentives such as extra-curricular credits and a point-based system within the app were implemented with the aim of incentivize sustained involvement and fostered a competitive environment through the school-wide leaderboard and post-game rewards. Thanks to this comprehensive approach, over 1,000 students (a quarter of WKU’s enrollment in 2023) responded to be interested in the WKU game. As expected, a fraction of those students dropped eventually, resulting in *N_total_* = 794 students participating in the WKU game. A separate manuscript currently in preparation delves into the methodological aspects of the WKU field behavioral experiment, providing in-depth details on how it was planned, promoted, and carried out by a team of WKU students [45].

**Figure 2:**
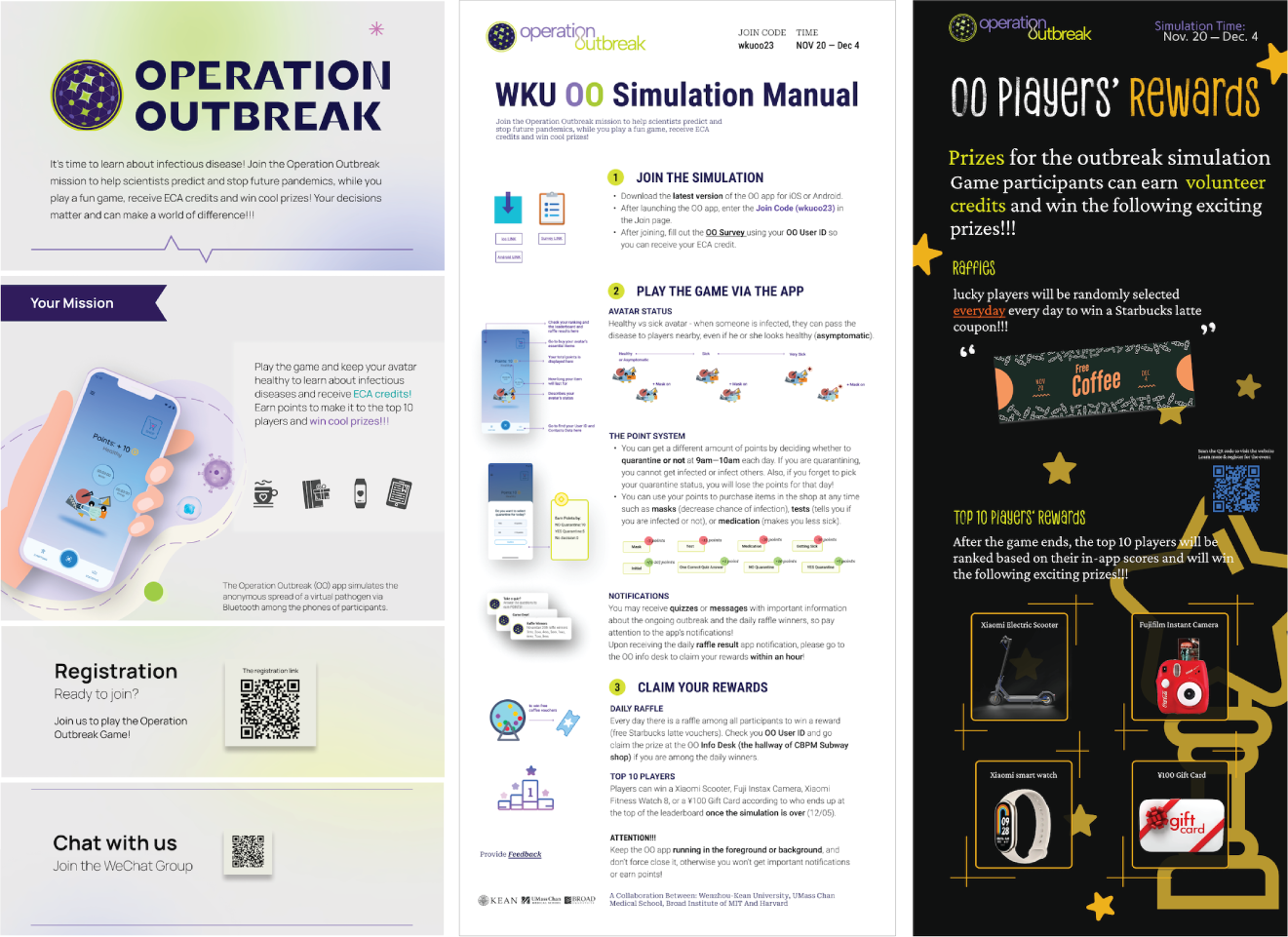
Materials used to promote the experimental epidemic game at WKU and facilitate student enrollment into the activity. Section of an informational micro-site about WKU game with links to the registration form and WeChat group (left). Manual page detailing the use of the app and the features specific to the WKU game (center), and flier describing the final prizes to be awarded to top-scoring students (right).

### 2.2 Statistical analysis

The data were, first of all, analyzed using a simple statistical approach to understand the patterns of the experiment’s dynamics. Fig. 3 (a) displays the average number of infections per hour, revealing specific times of day with heightened transmission activity. Fig. 3 (b) presents the daily total infections (brown line) along with a 7-day moving average (purple line), which smooths out short-term fluctuations to highlight longer-term trends. Fig. 3 (c) shows the estimated effective reproduction number (*R_eff_*), with gray points representing the estimated values, error bars indicating the 95% confidence intervals, and a smoothed trend black line, offering insights into the transmission potential over time. The red dashed line at *R_eff_* = 1 serves as a critical threshold, indicating whether the infection is spreading (if *R_eff_ >* 1) or declining (if *R_eff_ <* 1). We used the serial interval information for COVID-19 from China as a proxy for the serial interval of the app-based outbreak, properly scaled to account for the two-week duration of the experiment [46]. These data provide view of infection dynamics, peak transmission periods, and the impact of interventions on outbreak control.

**Figure 3:**
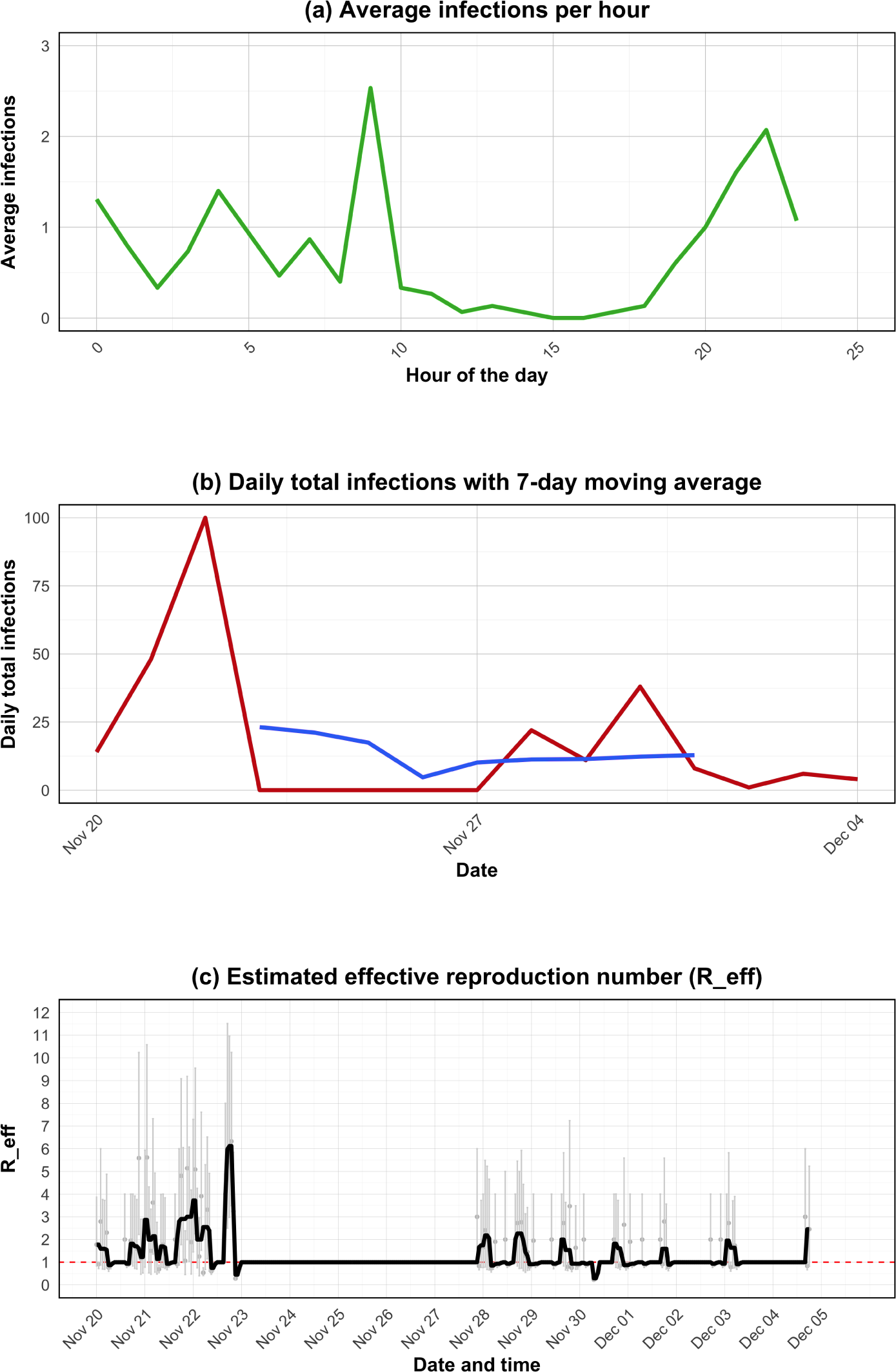
Extended time series analysis of infection dynamics during the WKU game (a) Hourly average number of infections, highlighting periods of heightened transmission activity throughout the day. (b) Daily total infections (red line) accompanied by a 7-day moving average (blue line) to smooth short-term fluctuations and reveal longer-term trends. (c) Estimated effective reproduction number (*R_eff_*), where gray points represent the estimated Reff values, error bars denote the 95% confidence intervals, and the black line indicates the smoothed trend. The red dashed line at *R_eff_* = 1 marks the critical threshold, above which the infection is spreading and below which it is declining.

Even though network epidemiology is not the methodological approach we followed in this manuscript, we conducted some basic statistical analyses on the contact network and transmission tree that resulted from the WKU game. First of all, these analyses revealed that a fraction of the participants (198) did not seemingly make any contacts with anybody else. Upon closer examination, we discovered that most of those participants were using a brand of Android smartphones that was not compatible with the Bluetooth library in the app. Therefore, we removed those 198 participants, and further removed 122 participants who had interacted for less than 5 minutes in total during the 14 days of the WKU game and 2 participants who formed an isolated dyad. We used the remaining *N_network_* = 472 = 794 − 198 − 122 − 2 in all subsequent network analyses (see Fig A4). We applied Taube et al. [47] methodology that characterizes the superspreader epidemiology in the transmission tree of any real outbreak by calculating two tree statistics, the proportion of cases causing super spreading events and the dispersion parameter. In the case of the WKU data (Fig A4), we found that it closely mirrors patterns seen in outbreaks from biological pathogens (refer to the Appendix for details). This result provides support for the external validity of the data from WKU game.

Survey responses captured basic demographics of the participating students, and their perceptions of quarantine/isolation benefits and costs, adapted from H1N1 pandemic research [50]. Detailed daily counts of within-app quarantine, isolation, mask purchases, and rapid diagnostic test use provided a detailed view of decision-making during the epidemic game. Note that only participants who downloaded the app and registered were included in the analysis, as the app tracked interactions solely among active participants.

The appendix provides a summary of the survey responses, which were not used in our present analyses, but we are planning to incorporate in follow-up work to this manuscript. This comprehensive dataset is a valuable resource for epidemiological research, health policy evaluation, and public health, enhancing our understanding of outbreak dynamics and human behavioral responses [3, 48, 49, 51, 52], and it is available on a Zenodo public repository (see Declarations section for the repository URL.)

### 2.3 Conceptual model framework

We employed an SEIR (Susceptible-Exposed-Infectious-Recovered) model framework [53, 54] to investigate pathogen transmission dynamics within a controlled university environment, specifically WKU through the experimentation platform and the app; see full model in Fig. 4. Our model incorporates multiple transmission pathways, accounting for both asymptomatic and symptomatic exposure, and captures the quarantine/isolation statuses of individuals. The population is assumed to be constant in size and homogeneous in structure, assumptions that are justified given the two-week duration of the experiment during a class semester and the residential nature of the WKU campus. Beyond these simplifying assumptions, the model allows for variations in vulnerability and transmissibility, thus reflecting the complex and heterogeneous nature of real-world disease spread.

**Figure 4:**
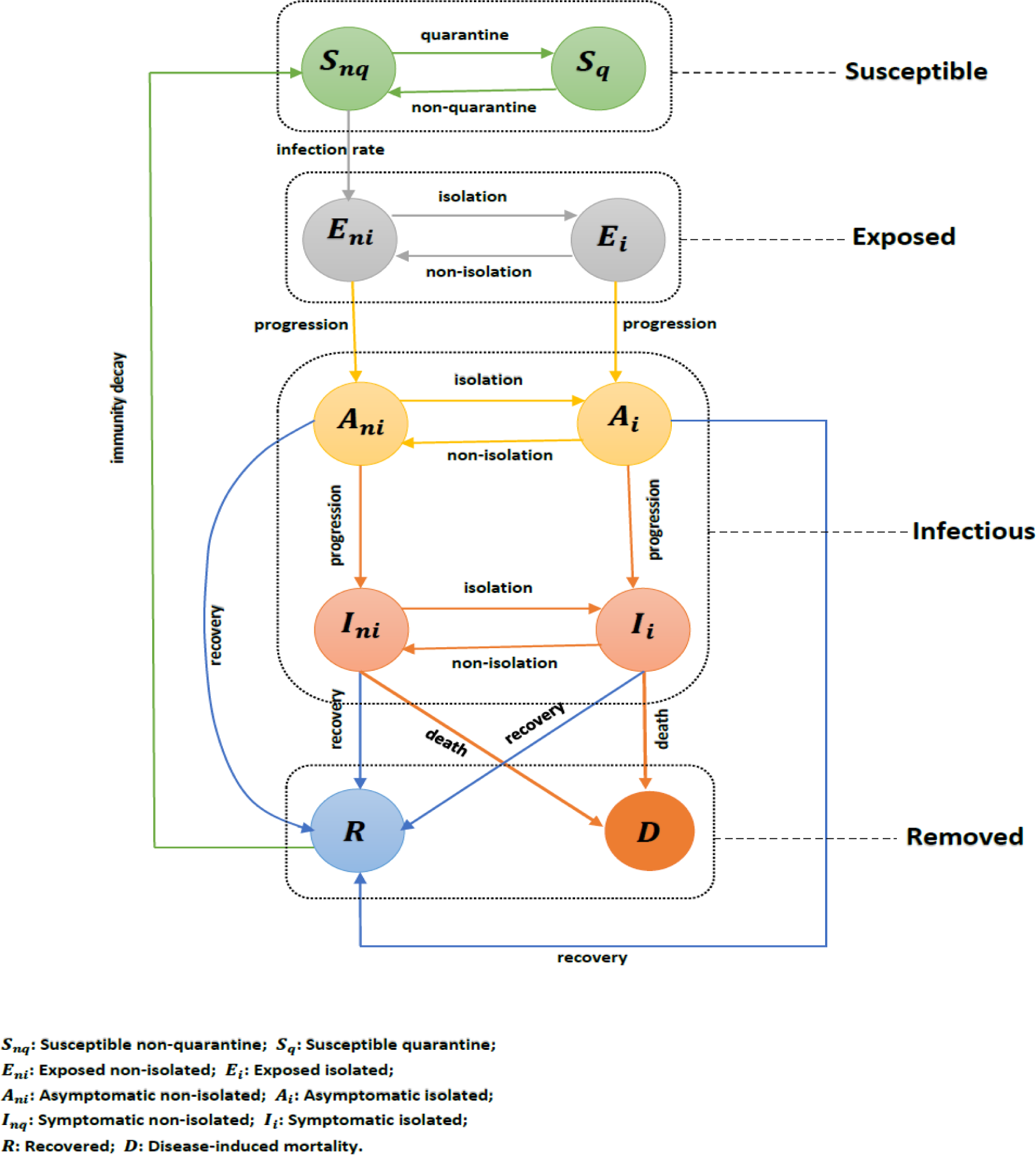
Flow chart for the model defined by equation (1) for pathogen transmission dynamics within the WKU setting. Solid arrows indicate the transitions between compartments, with the associated *per capita* flow rates shown next to each arrow. The model partitions the total population into ten compartments: non-quarantine susceptible (*S_nq_*), quarantine susceptible (*S_q_*), non-quarantine exposed (*E_ni_*), isolated exposed (*E_i_*), non-isolated asymptomatically infected (*A_ni_*), isolated asymptomatically infected (*A_i_*), non-isolated symptomatically infected (*I_ni_*), isolated symptomatically infected (*I_i_*), recovered (*R*), and deceased (*D*) individuals. The state variables and parameters are defined in Table A1, and parameters values in Table A2.

A key feature of our model is its integration of human behavior as a dynamic factor that influences transmission rates. We assume that during the experimental game, participants’ adherence to NPIs—such as mask-wearing, social distancing, and isolation—is driven by their perception of infection risk, which evolves in response to changes in infection severity and prevalence within the simulated environment. This feedback loop between behavior and transmission dynamics is crucial for accurately modeling the spread of infectious diseases in real-world scenarios where human decisions play a pivotal role. It is important to distinguish this post-experiment SEIR model with human behavior feedback from the model used by the app to drive the transmission during the experiment. We fit the parameters of the former using only the data gathered from the experiment, and we set the “ground-truth” parameters of the latter beforehand to ensure that the experimental game would take place within the desired time and case-count bounds.

Our app platform provides a unique experimental environment that generates rich datasets, including detailed records of contact tracing, participant behaviors, quarantine/isolation status, infection events, and (simulated) disease outcomes. By leveraging these data, we were able to construct a model to quantify the impact of various intervention strategies, such as contact tracing protocols, quarantine/isolation measures, and NPIs, on pathogen transmission. The model’s flexibility allows us to represent different NPIs mathematically, capturing how these interventions collectively reduce transmission. Additionally, our framework considers the dynamic nature of behavior, where individuals can alter their adherence to NPIs based on perceived risks and evolving conditions within the proximity-based experimental game. This approach provides a comprehensive tool for assessing the effectiveness of public health interventions and informs strategies for outbreak management and pandemic preparedness. The insights gained from integrating the experimental data into our SEIR model not only advance theoretical understanding but also offer practical guidance for controlling infectious disease outbreaks in educational settings and beyond.

### 2.4 Model simplification

We used a simulation-based inference framework for epidemiological dynamics to fit our model to the experimental data, as proposed by [59]. The analysis focuses on a specific scenario of model (1) within a closed domain (WKU campus). To streamline the model, we used coupling dynamics by combining quarantine/isolation and non-quarantine/non-isolation compartments due to their similar dynamic nature (except for the constant and flexible transmission rate) [83].

This simplification is justified by the analogous behavior of the full and sub-models, as well as the ability to capture human behavior and NPIs through a flexible transmission rate described in equation (1). Consequently, the simplified model retains the same incidence function and flexible transmission rate, with varying parameters *P* and *C* representing risk perception from participants and cumulative cases, respectively, which are key in changing the transmission behavior. This approach allows us to assess the influence of human behavior and intervention strategies on pathogen transmission during the WKU game. By analyzing data from the experiment, we can evaluate the effectiveness of different control measures and understand the role of human behavior and attitudes in mitigating outbreaks. Furthermore, numerical assessments and sensitivity analyses of the model’s parameters provide insights into the robustness of our findings and model assumptions. Therefore, the simplified version of the model (depicted in Fig. A6), which maintains the same incidence function as in the full-model, while using flexible transmission rate, is given by:

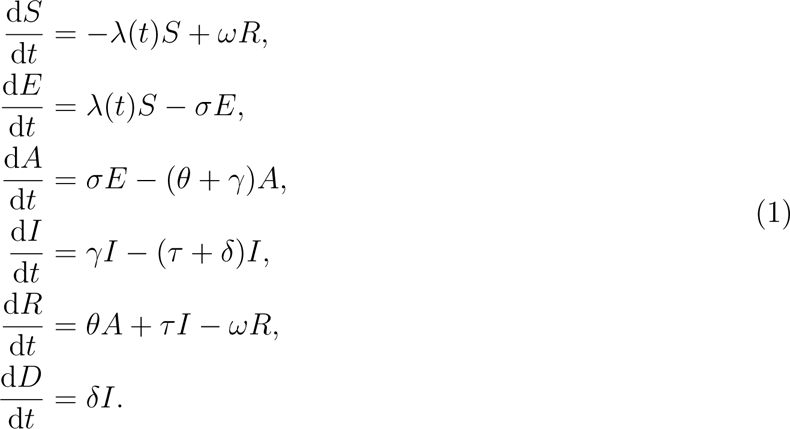

The sub-model (1) was augmented with an time-varying variables, “*P* “, measuring the participants’ perception of risk regarding the number of symptomatic cases, so that

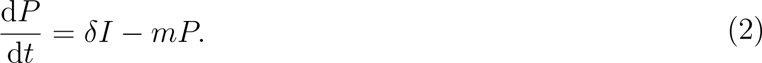

where *m* represents the mean duration of participants’ reaction to the number of infections.

In addition, we used a time-varying transmission rate introduced by He et al. [56], which is also known as effective (flexible) transmission rate, denoted by *β*(*t*). It accounts for intervention actions (modeled as a step function) and the reduction in contacts among individuals in response to the proportion of cases, reflecting the epidemic’s severity. This variable transmission rate *β*(*t*) is formulated in equation (3) below

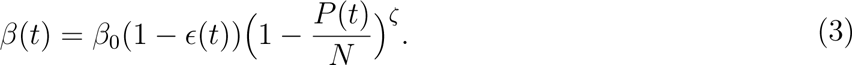

Here, the parameter *ɛ* represents interventions’ strength and *ζ* aims to capture individuals’ response intensity. Within this framework, the parameter *P* denotes risk perception. The individuals’ perception of risk increases with the number of infections, while decreasing over time, as reflected by the two terms in equation (2). The parameters *β* and *ɛ* are stepwise functions of time.

The dynamics between intervention strength (*ɛ*), response intensity (*ζ*), and risk perception (*P*) are critical for understanding and predicting disease spread. Adaptive behavioral responses, influenced by real-time changes in perceived risk, significantly shape epidemic trajectories, and our parameter choices can be justified as follows:

- **Intervention Strength (***ɛ*): Represents the impact of public health measures, as the result of their intrinsic effectivity but modulated by disease severity and compliance. Higher perceived severity typically increases *ɛ*, as it induces compliance with preventive actions.
- **Response Intensity (***ζ*): Measures variability in individual responses to perceived infection risk, often influenced by cultural and personal factors. As *ζ* increases with social awareness and effective communication, disease transmission would decrease from a baseline given by the level of risk perception in the population.
- **Risk Perception (***P*): Individuals’ perception of risk can respond to many factors, from pathogen infectivity, information access, and pre-existing attitudes. Heightened *P* can boost preventive adherence but may also lead to panic or misinformation, complicating control efforts.

Obviously, the relationship between these three parameters and their effect on disease transmission is potentially very complex. We have reached a tractable model by adopting equations (2) and (3) making the flexible transmission rate dependent on *ɛ*, *ζ*, and *P*. In turn, *P*, with its dependence on the number of infections, *I*, and a built-in progressive decrease reflecting peoples’ physiological traits such as forgetfulness and normalization, introduces these additional important elements into the transmission dynamics. Although simple, this framework has been applied extensively to model real outbreaks with satisfactory results [39, 55, 56, 85, 86].

Note that individuals in quarantine and isolation are restricted from further spreading the disease. As such, they do not contribute to transmission, and therefore, *λ*(*t*) in this case is defined as 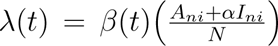, where *α* serves as the modification parameter representing the reduced infectiousness. Also, our model assumes a Poisson process due to the low frequency of (simulated) fatal cases, which aligns well with the Poisson distribution’s suitability for rare events. This approach is standard in epidemiological modeling for low-frequency events (see references [63, 71] for similar applications).

The mechanism of time-varying flexible transmission rate allows for the inclusion of human behavioral components, which have a considerable impact on disease dynamics. Specifically, we adopted a mechanistic transmission rate function based on prior studies [55, 56] to include individual behavioral actions. We consider three factors: (i) a time-varying risk perception by individuals (fear of infection, severe infection), modeled by *P* (*t*), (ii) level of adherence of individuals to NPIs such as self-isolation, quarantine, or reporting, modeled by the parameter *ζ*, and (iii) the effectiveness of the NPIs, modeled by the parameter *ɛ* (Refer to the equation (3) above.) This technique has been used in prior studies on infectious disease transmission, including COVID-19 [55] and influenza [56], and is adaptable to our experimental scenarios. We explored each mechanism by treating the key parameters as a flexible (cubic spline) or time-varying function and comparing various formulations’ fitting performance. Additionally, we can enhance the models in the future by incorporating further details, such as age structure, when data is available, to conduct more fine-grained experiments that could aid in pandemic preparedness and response. For further details and analysis of the model, see the Appendix section.

## 3 Results: Evaluating the impact of individual perception and NPI strength

### 3.1 Influence of individual perception and NPIs

Our experimental data results provide critical insights into the dynamics of pathogen transmission under varying levels of individual perception and NPIs during the WKU game. Fig. 5 illustrates the daily new cases across three distinct scenarios: a naive scenario with no interventions (*ɛ* = 0 and *ζ* = 0), a scenario driven by individual perception (only *ɛ* = 0), and a combined scenario incorporating both individual perceptions and NPIs. In the naive scenario, represented by the orange solid curve, a rapid increase in new cases is observed, highlighting the unmitigated spread of the pathogen in the absence of any intervention. This serves as a baseline, demonstrating the potential severity of an outbreak without any behavioral change or NPIs. The second scenario, depicted by the red dashed curve, models the impact of individual behavioral responses to the perceived risk of infection. Here, participants adjusted their behavior based on the evolving outbreak situation, such as by reducing contacts or increasing protective measures like mask-wearing. The substantial reduction in new cases in this scenario underscores the significant role that individual perception plays in controlling disease spread. This finding is consistent with previous research that emphasizes the importance of individual actions in epidemic management [55, 56]. The third scenario, represented by the green solid curve, combines individual perception with structured NPIs, such as quarantine and isolation. This scenario shows the most pronounced decrease in daily new cases, illustrating the enhanced effectiveness of combining adaptive human behavior with coordinated public health interventions. The grey curve with dots, representing simulated reported cases, aligns closely with the observed data, further validating the accuracy and predictive capability of our model.

**Figure 5:**
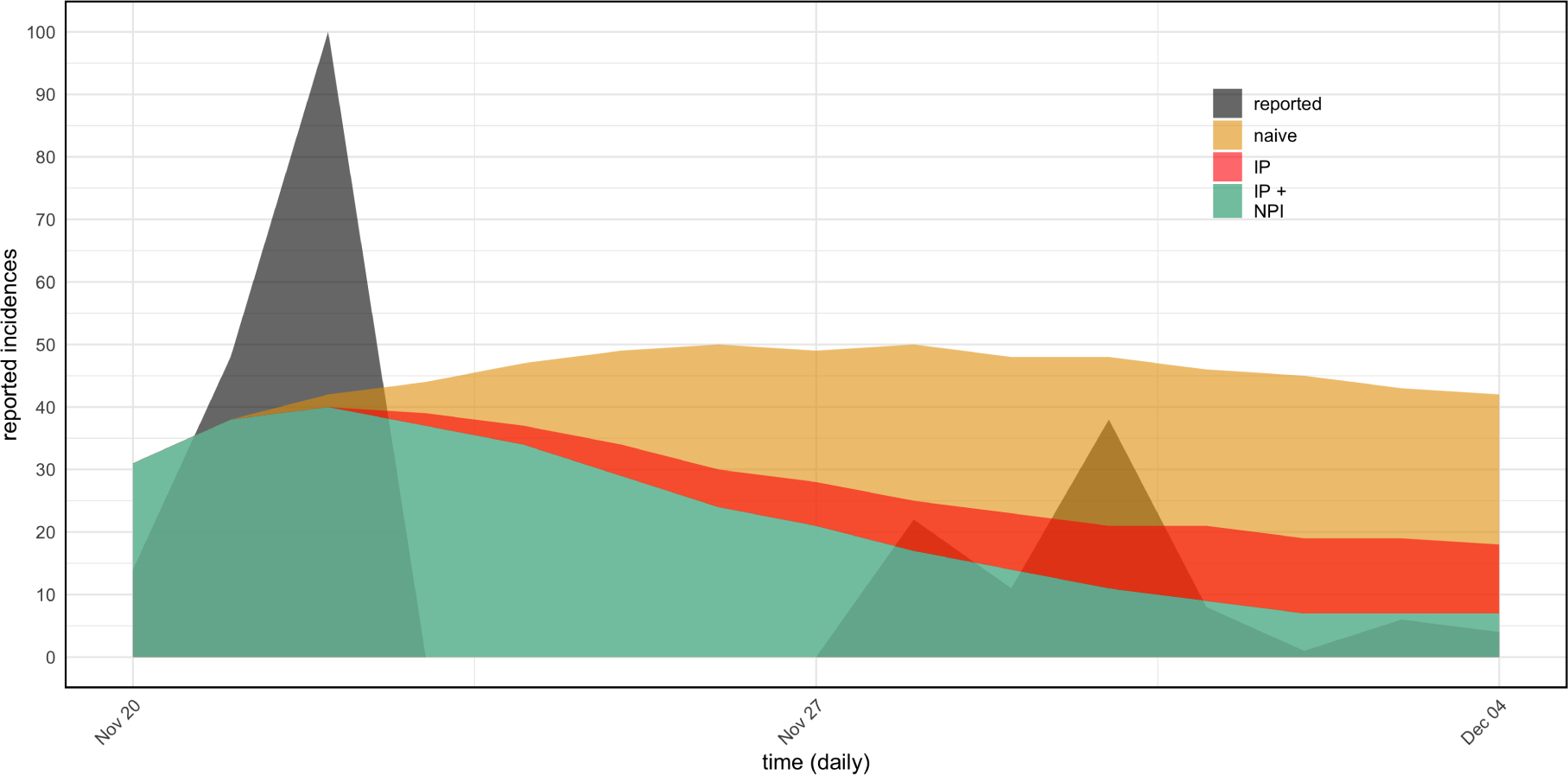
Daily new cases across three intervention scenarios during the WKU game represented by the black shadowed curve. The orange solid curve represents the naive scenario of the simulation with no interventions, showing a rapid increase in new cases. The red curve illustrates the scenario with individual perception based on behavior change, resulting in significant reduction in new cases. The green solid curve depicts the scenario combining individual reactions with NPIs such as quarantine and isolation, showing the most pronounced decrease in daily new cases. The grey curve with dots represents the simulated reported cases, aligned with official data, validating the accuracy of our model. Effect of intervention take some time (a couple of days) to show up in the data.

### 3.2 Evaluation of reporting ratios and model validation

Fig. 6 (a) & (b) presents the reporting ratios between observed cases and model estimates under two scenarios: (a) the combined scenarios of individual perception and NPIs and (b) the scenario with individual perception only. The relatively stable reporting ratio observed in both scenarios indicates that the model effectively captures the dynamics of reporting behavior and the efficacy of interventions. This stability is crucial for ensuring that the model accurately reflects real-world dynamics, thereby providing reliable predictions of outbreak progression. The consistent reporting ratios also underscore the effectiveness of combining individual behavioral responses with structured NPIs in mitigating pathogen spread and ensuring reliable reporting. These findings highlight the importance of coordinated interventions that integrate both human behavior and public health strategies to achieve optimal outcomes during an epidemic. The insights gained from these simulations results set the stage for a detailed sensitivity analysis, which will be discussed in the subsequent section. This analysis aims to identify key parameters that drive transmission dynamics and intervention effectiveness, offering further guidance for optimizing public health strategies during infectious disease outbreaks.

**Figure 6:**
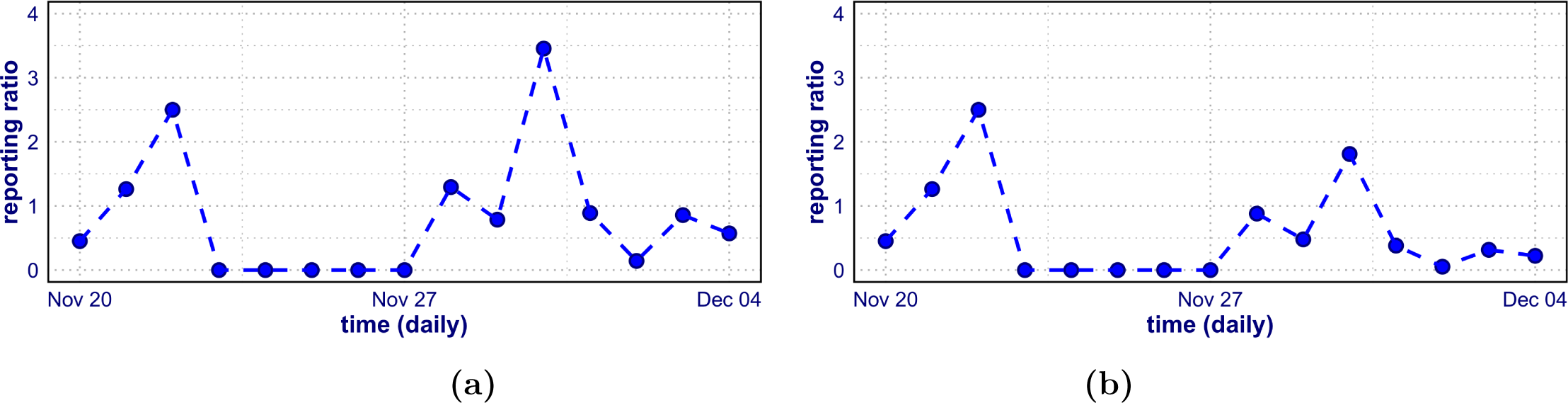
Reporting ratio between observed cases and model estimates under the combined scenario of individual perception (IP) and non-pharmaceutical Intervention (NPIs) (a) and IP only (b), respectively. The relatively stable ratio indicates that the model accurately captures the dynamics of reporting behavior and intervention efficacy. This stability underscores the effectiveness of coordinated interventions in mitigating the pathogen’s spread and ensuring consistent reporting accuracy.

### 3.3 Sensitivity analysis

We conducted sensitivity analyses to assess the influence of our model’s parameters on the transmission trajectory. The analysis underlined the varying relevance of each parameter and offered a more profound understanding of the underlying dynamics. The findings emphasize the critical role of prompt intervention and adherence to preventive measures in reducing the severity of the outbreak.

#### 3.3.1 Global sensitivity analysis

In this sub-section, we aim to provide a paradigm that takes into account both individuals’ perception of risk (which is modelled by *ζ*) and NPIs’ strength (which is modelled by *ɛ*), as well as a time-dependent reporting rate through sensitivity analysis, and also to determine how the parameters *ζ* and *ɛ* affect the pathogen spread [55]. Our analysis demonstrates that both *ζ* and *ɛ* play vital roles in shaping transmission patterns, although *ζ* shows greater magnitude. Specifically, we found that *ζ* exerts a more pronounced effect on transmission dynamics compared to *ɛ*. Small increases in *ɛ* or *ζ* could result in decreased transmission, as shown in Fig. 7 (a) & (b), but the impact is relatively low for *ɛ* compared to *ζ*, which causes substantial decreases in transmission, indicating a higher sensitivity to changes in this parameter. This result stresses the role of *ɛ* and *ζ* in determining the course of an epidemic and highlights the importance of tailored measures to effectively control outbreaks. Therefore, through sensitivity analysis, we have gained valuable insights into the relative influence of *ɛ* and *ζ*, enabling us to optimize intervention strategies and enhance our understanding of the underlying transmission mechanisms. This knowledge is crucial for informing public health policies and improving pandemic preparedness.

**Figure 7:**
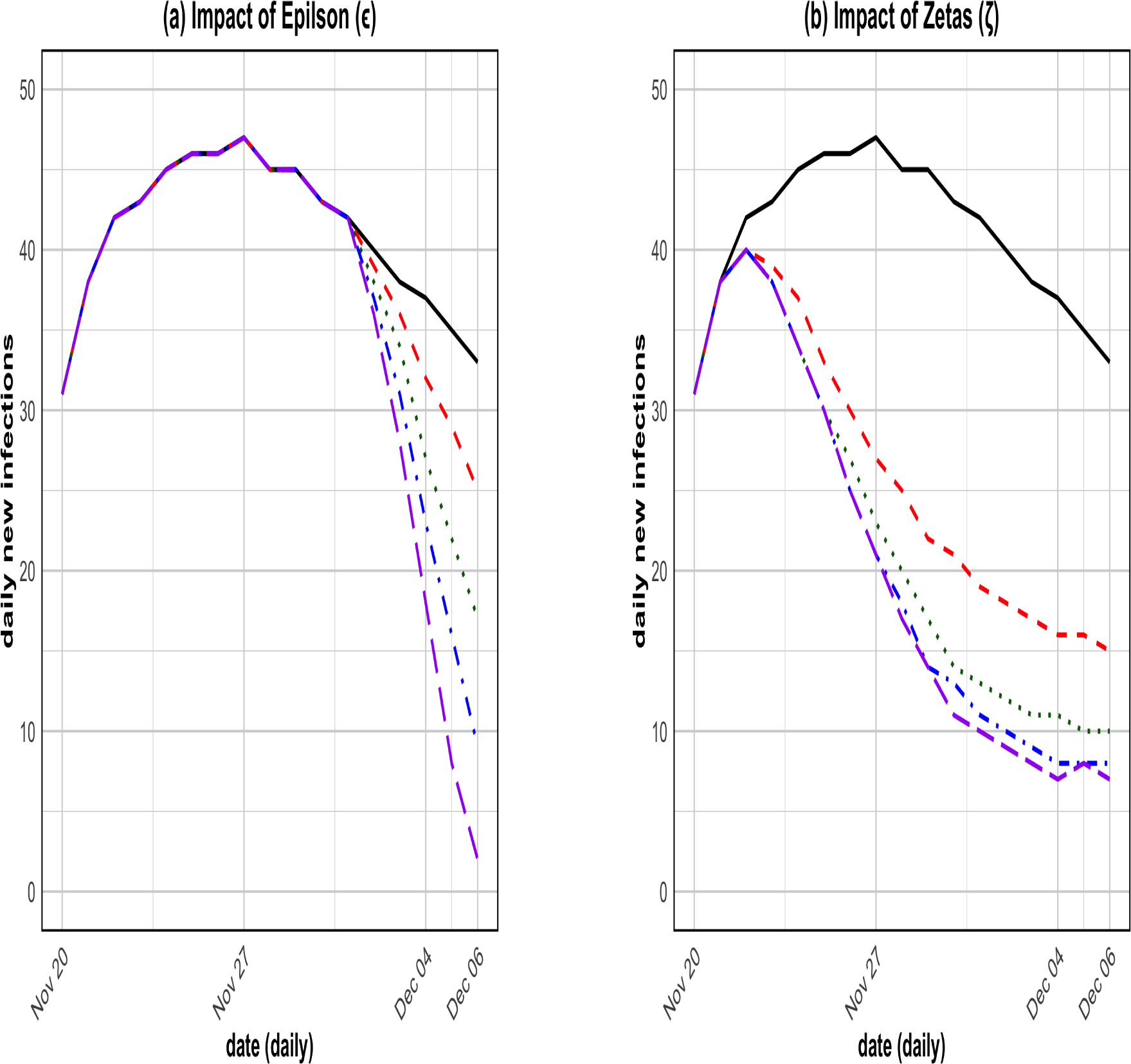
Sensitivity analysis illustrating the effects of parameters ɛ (a) and ζ (b) on transmission dynamics. Our analysis reveals that while both parameters significantly influence the epidemic trajectory, ζ (which measures the intensity of individual response) has a notably stronger impact than ɛ (which measures the intervention strength), which indicates that modest increases in ɛ result in a measurable reduction in transmission, but even a slight increase in ζ leads to a much more substantial decrease in transmission rates. This highlights the critical role of ζ in effective pathogen control, surpassing the influence of ɛ.

#### 3.3.2 Local sensitivity analysis

To delve deeper into our model’s dynamics and parameter effects, we conducted a sensitivity analysis using partial rank correlation coefficients (PRCCs) to assess how model parameters influence the overall transmission [62, 67]. Utilizing PRCCs with *R*_0_ as the response function, we identified the most effective parameters influencing the spread of the pathogen. See Fig. 8(a) and (b). Our sensitivity analysis involved sampling 5,000 random instances from uniform distributions within the specified parameter ranges. Each instance was simulated to produce biological outcomes, and PRCCs were calculated between parameters and biological values. The PRCCs for *R*_0_ highlighted that the transition rate of exposed non-isolated individuals to the infected non-isolated class, along with the probability of transmission per contact, significantly influence *R*_0_. These findings emphasize the importance of targeted interventions to reduce interactions among susceptible and infected individuals and decrease the probability of transmission per contact. Even minor adjustments to these critical parameters could substantially impact the infection’s spread, underscoring the necessity for effective public health measures and timely interventions to control the outbreak.

**Figure 8:**
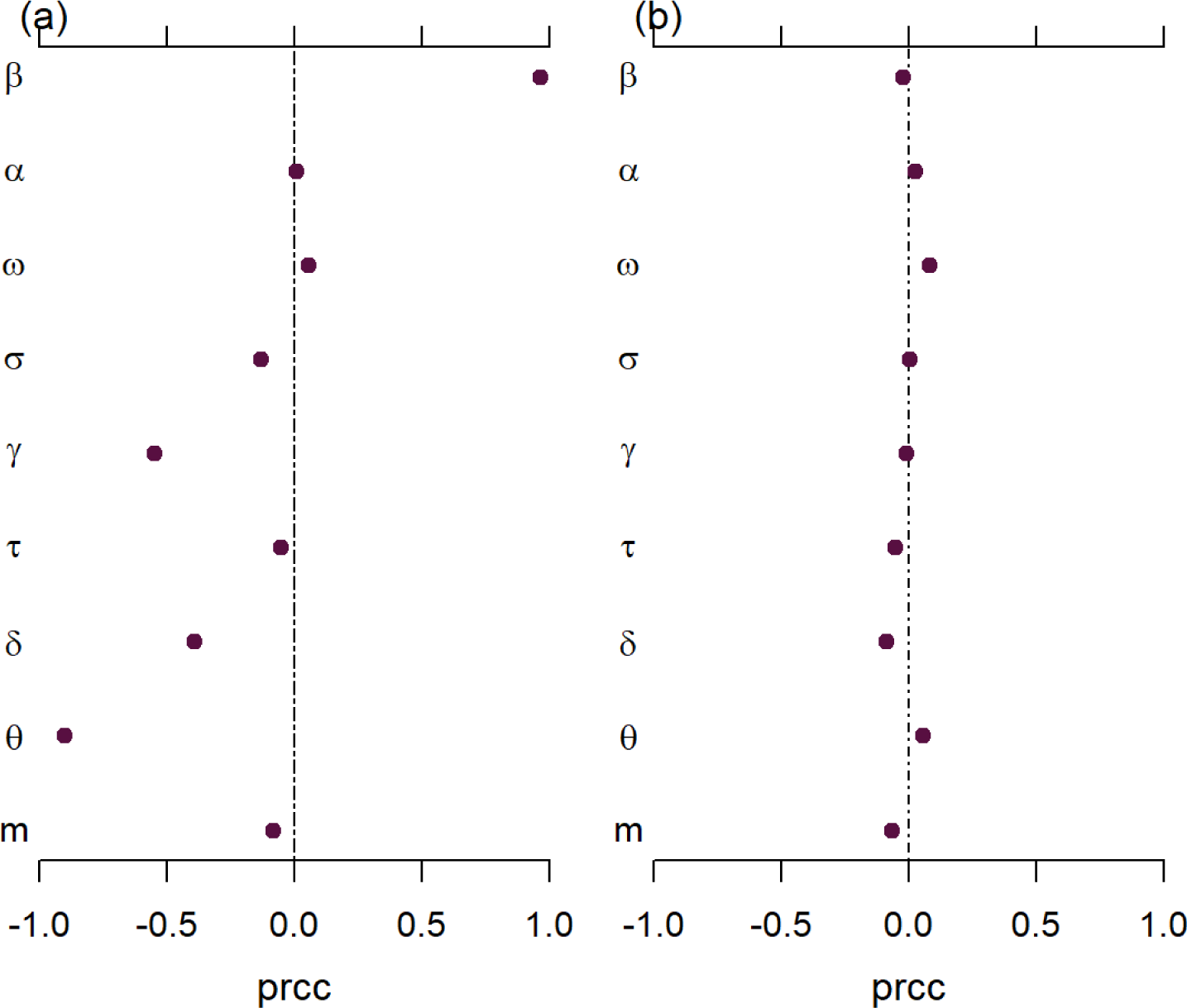
Local sensitivity analysis using (a) R0 and (b) the attack rate as response functions, assessed through partial rank correlation coefficients (PRCCs). The analysis identifies the transition rate from exposed non-isolated individuals to the infected non-isolated class, along with the probability of transmission per contact, as the most influential parameters on both R0 and the attack rate. These results emphasize the critical need for targeted interventions focusing on these key parameters to effectively control the outbreak and reduce transmission rates.

### 3.4 Numerical simulations

In this section, we conducted numerical simulations of the simplified model to evaluate the influence of key parameters on transmission dynamics. The contour plots illustrate the dependency of the basic reproduction number (*R*_0_) on several critical parameters. Fig. 9 (a) shows the relationship between *R*_0_, *β* (probability of transmission per contact), and *θ* (recovery rate of symptomatically infected individuals). The plot demonstrates that higher values of *β* result in an increase in *R*_0_, while higher *θ* values lead to a decrease in *R*_0_, underscoring the critical importance of reducing transmission and increasing recovery rates to effectively control pathogen spread. Fig. 9 (b) presents the relationship between *R*_0_, *δ* (disease-induced death rate), and *α* (modification parameter for decreased infectiousness). The results show that increasing both *δ* and *α* significantly lowers *R*_0_, highlighting the importance of interventions that reduce infectiousness and disease severity in mitigating outbreaks. Finally, Fig. 9 (c) depicts the relationship between *R*_0_, *τ* (recovery rate of symptomatically infected individuals), and *α* (modification parameter for decreased infectiousness). The plot reveals that higher values of *τ* and *α* both contribute to a reduction in *R*_0_, reinforcing the importance of rapid recovery and lowering infectiousness as effective strategies for outbreak control. These findings emphasize the critical role of reducing transmission probability, increasing recovery rates, and mitigating disease severity in controlling infectious disease outbreaks. While other parameters could also be explored in future simulations, we focus on *β*, *θ*, *δ*, *α*, and *τ* due to their significant influence on transmission dynamics in the model. These key parameters offer valuable insights into how targeted interventions can alter the course of an epidemic, demonstrating the robustness and applicability of our model to real-world public health scenarios.

**Figure 9:**
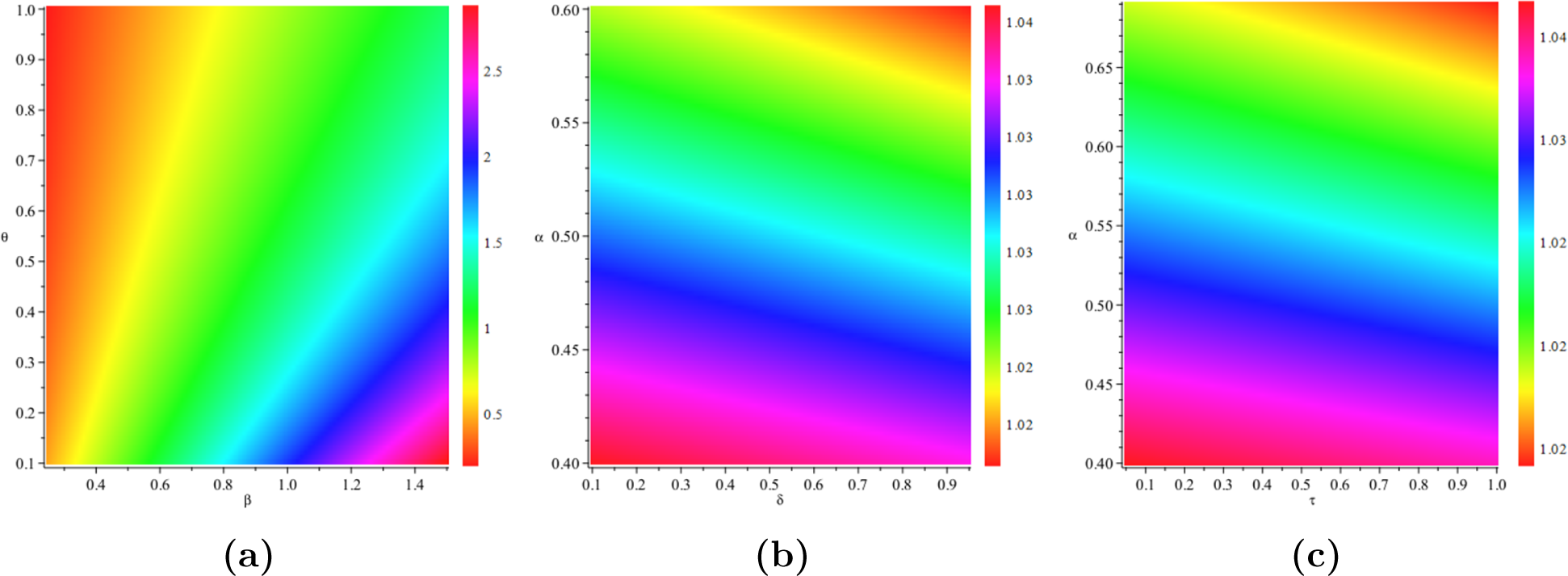
Contour plots of the basic reproduction number (R0) for the model (1), as a function of (a) transmission rate (β0) and recovery rate of symptomatically infected individuals (θ), (b) disease-induced death rate (δ) and modification parameter for decreased infectiousness (α), and (c) recovery rate of symptomatically infected individuals (τ) and modification parameter for decreased infectiousness (α), respectively. Parameter values used are provided in Table A2.

## 4 Discussion

The epidemic game conducted at WKU provided a unique opportunity to pilot our app-based experimentation platform during an open-world realistic “mock” outbreak, engaging nearly 1,000 students over two weeks. This large-scale proximity-based experiment integrated behavioral experiments and generated multimodal data, including contact and transmission patterns (see Figs. 3 and A1), highlighting the platform’s potential for interdisciplinary research. WKU students gained practical experience by participating in the experiment, while a subgroup of the students were actively involved in planning and organizing the activity around campus. The resulting data are available for instructors to use during classes and further epidemiological analyses. This data are particularly significant given its realistic representation of a real-life outbreak scenario in a university campus, thereby supporting the platform’s capacity to generate rich multimodal epidemiology datasets with external validity. [3, 22, 25].

Our in-depth study used a robust modeling approach to investigate pathogen spread during the WKU game. We designed an effective model framework (Figs. 4 & A6) to assess intervention strategies including quarantine and isolation, incorporating human behavioral perspectives and non-pharmaceutical interventions (NPIs). We conducted detailed model simulations and sensitivity analyses using the data generated from the experiment. Model fitting and parameter estimates (Fig. A7) demonstrated that our flexible transmission rate model effectively describes the transmission dynamics observed in the experiment. The inclusion of human behavioral factors, such as individual adherence to preventative actions and the assessment of risk, greatly enhanced the model’s accuracy. The predicted parameters accurately depicted the transmission situations, highlighting the significance of prompt and synchronized interventions [3, 43, 56].

Our model represents a significant step forward in the integration of dynamic behavioral factors into epidemiological modeling. Unlike traditional models that often treat behavioral responses as static or oversimplified, our approach accounts for the fluidity of human behavior during an outbreak. Individuals in our model can adjust their adherence to NPIs such as social distancing, mask-wearing, isolation and quarantine based on real-time changes in disease severity and perceived personal risk [30, 36]. This dynamic response is critical for understanding the true impact of public health interventions and for predicting the course of an epidemic [37, 38].

However, while our model incorporates a more realistic representation of behavior, it does not fully capture the entire spectrum of potential responses. Behavioral heterogeneity, influenced by factors such as cultural norms, misinformation, and varying levels of trust in public health messages, presents a complex challenge for modeling [39, 40]. Our findings suggest that while the inclusion of dynamic behavior significantly improves the accuracy of epidemiological predictions, there is still a need for further refinement. Future models should aim to incorporate more granular data on behavioral responses, potentially integrating psychological and sociological insights to better capture the diversity of human actions in the face of an epidemic. This ongoing refinement will be crucial for developing models that can more effectively inform public health strategies and interventions, particularly in diverse and rapidly changing real-world settings [30, 36].

The model simulation results presented in Fig. 5 underscore the profound impact of individual behavior and NPIs on disease transmission dynamics. The naive scenario, which operates under the assumption of no interventions, illustrated a rapid and uncontrolled spread of the pathogen, emphasizing the critical need for timely interventions to prevent widespread transmission. In contrast, scenarios incorporating individual reactions to the outbreak demonstrated a significant reduction in new cases. The red dashed curve in Fig. 5 reflects this impact, where behavioral responses such as social distancing, mask-wearing, and self-isolation markedly decrease transmission rates. Human behavior plays a pivotal role in controlling the spread of infectious diseases, as individuals’ decisions to adopt preventive measures can substantially alter the trajectory of an epidemic [55, 56, 44]. The most pronounced decrease in daily new cases was observed in the scenario combining individual reactions with NPIs, depicted by the green solid curve. This scenario incorporates measures such as quarantine, isolation, and contact tracing alongside individual behavioral responses. The synergistic effect of these combined strategies results in a substantially flatter epidemic curve, demonstrating the efficacy of coordinated intervention efforts. The alignment of simulated reported cases with official data, shown by the grey curve with dots, further validates the model’s accuracy and predictive capability.

The analysis of the reporting ratio in Fig. 6 adds another layer of validation to our model. The stability of the reporting ratio when both individual reactions and NPIs were implemented indicates that the model accurately captures the dynamics of reporting behavior and the effectiveness of interventions. This consistency is crucial for effective outbreak management, ensuring reliable data for decision-making and policy formulation. Accurate and stable reporting is essential for monitoring the outbreak and adjusting intervention strategies in real-time. These findings emphasize the critical role of NPIs and individual behavior in managing the spread of infectious diseases. Timely and decisive interventions, coupled with public compliance, are key to controlling outbreaks. Public health strategies must prioritize raising awareness about preventive measures and ensuring public cooperation. Campaigns to educate the public on the importance of these measures can significantly enhance compliance and effectiveness of interventions. The robustness of our model, validated through these simulations and sensitivity analyses, further underscores its applicability in real-world epidemiological studies and public health planning. By integrating human behavior and NPIs, the model provides a comprehensive tool for predicting outbreak trajectories and designing effective intervention strategies. This integrated approach is essential for optimizing public health responses and improving pandemic preparedness [25, 65].

Our study aligns with previous research indicating that combined individual and population-level actions are crucial in mitigating the spread of infectious diseases [25, 55, 56]. The insights gained from this experiment provide valuable guidance for future outbreak management and emphasize the importance of coordinated, multifaceted intervention strategies [25, 69]. Furthermore, our model simulation results demonstrate that individual behavior and NPIs are indispensable in controlling disease transmission. The effectiveness of these measures in reducing new cases and maintaining stable reporting ratios highlights the importance of a comprehensive and coordinated public health response. These findings provide a robust foundation for developing strategies to manage future outbreaks, ensuring both public compliance and effective intervention implementation.

To further highlight the effectiveness of key parameters such as *ɛ* (which measures the efficacy of intervention) and *ζ* (which measures the intensity of behavioral response), global sensitivity analysis simulations were conducted and depicted in Fig. 7. Our findings showed that minor increases in *ɛ* and *ζ* can significantly reduce transmission, with *ζ* having a more noticeable impact. This emphasizes the significance of robust and intensive measures in containing the outbreak. In addition, local sensitivity analysis (see Fig. 8), which has *R*_0_ as the response function, analyzed based on the PRCC, identified the transition rate of exposed non-isolated individuals to the infected non-isolated class (*σ_n_*) and the probability of transmission per contact (*β*) as the most critical parameters impacting *R*_0_. These variables have a major impact on basic reproduction numbers, indicating their significance in control. Furthermore, contour plot simulations provided valuable visual insights into the interactions between critical model parameters and their impact on the basic reproduction number, *R*_0_. In Fig. 9 (a), the simulations highlight the relationship between *β* (probability of transmission per contact) and *θ* (recovery rate of symptomatically infected individuals), revealing that higher values of *β* significantly increase *R*_0_, while increasing *θ* effectively reduces *R*_0_. This underscores the importance of both limiting transmission rates and enhancing recovery efforts in mitigating the spread of the pathogen. Similarly, Fig. 9 (b) illustrates the sensitivity of *R*_0_ to *δ* (disease-induced death rate) and *α* (modification parameter for decreased infectiousness). The simulations show that increases in both *δ* and *α* result in a notable reduction in *R*_0_, highlighting the critical role of interventions aimed at reducing disease severity and infectiousness in controlling outbreaks. Lastly, Fig. 9 (c) depicts the relationship between *R*_0_, *τ* (recovery rate of symptomatically infected individuals), and *α*. The results demonstrate that higher recovery rates and reduced infectiousness both contribute significantly to lowering *R*_0_, emphasizing the importance of rapid treatment and measures aimed at reducing the spread of infection. These contour plot simulations reinforce the notion that strategic interventions targeting key parameters—such as transmission probability, recovery rates, and infectiousness—are essential for effective epidemic control. By focusing on reducing transmission and enhancing recovery efforts, public health strategies can be optimized to limit the spread of disease and lower the reproduction number [25, 69]. This analysis underscores the critical need for coordinated, data-driven interventions tailored to the specific dynamics of each outbreak. Further numerical simulations (presented in A8) highlight the significant influence of key epidemiological parameters—transmission rate (*β*_0_), recovery rate (*θ*), and progression rate (*γ*)—on disease dynamics. Variations in *β* notably impact both asymptomatic and symptomatic infections, with a 50% increase leading to substantial rises in case numbers, underscoring the need for interventions like social distancing and mask-wearing to mitigate transmission. Similarly, enhancing recovery rates (*θ*) by 50% reduces the infectious period for asymptomatic individuals, while slowing the progression from asymptomatic to symptomatic stages (*γ*) helps to alleviate the burden of symptomatic cases. These findings emphasize the critical role of reducing transmission, speeding up recovery, and delaying disease progression in controlling outbreaks effectively.

## Conclusion

In conclusion, this study elucidates the significance of our platform as a powerful tool for studying pathogen transmission and evaluating the impact of human behavior and NPIs on disease dynamics via an open-world experimental game approach. The integration of a flexible, time-varying transmission rate model in the analyses significantly enhances our understanding of how behavioral factors and NPIs influence the spread of infectious diseases. By accurately capturing these complex dynamics, our model provides critical insights into effective outbreak management strategies, underscoring the importance of timely, coordinated interventions and adherence to preventive measures. Our experimentation platform’s ability to generate realistic multimodal datasets—encompassing epidemiological, behavioral, and network data streams—could prove very valuable for public health planning and pandemic preparedness. The robustness and applicability of our model, validated through rigorous simulation and sensitivity analyses, highlight its potential as an important tool for guiding public health policies and optimizing response strategies in future outbreak scenarios. Future research will aim to refine this approach by incorporating additional data streams and elements such as age structure and varying interaction patterns, thereby advancing our understanding of disease dynamics. Coupling the model with a game-theoretic approach to analyze behavior during the experiments could further enrich our insights into outbreak management, ultimately contributing to more effective public health strategies and a deeper preparedness for future health crises.

### Strengths

Our study on the experimental game using the epidemic app at WKU demonstrates significant strengths. The large-scale, real-life experiment, involving nearly 1,000 students over two weeks, provided a detailed dataset closely mirroring real-world outbreaks. This rich dataset enabled rigorous analysis of pathogen transmission dynamics and the effectiveness of intervention strategies. A key strength of this research is the integration of behavioral data with epidemiological modeling. Our experimentation platform facilitated the collection of multimodal data, including epidemiological, behavioral, and network streams, offering a comprehensive view of disease dynamics. Our model, which incorporates human behavior and NPIs, accurately reflected the transmission patterns observed during the experiment. The robustness of our approach is further supported by extensive simulations and sensitivity analyses, validating the model’s accuracy and its applicability to public health planning. Moreover, the platform’s ability to generate realistic, high-quality data underscores its potential as a valuable tool for enhancing outbreak response strategies and pandemic preparedness.

### Limitations

Despite the strengths of our study, we must acknowledge several limitations. The assumption of homogeneous mixing within sub-populations may oversimplify real-world interactions, potentially affecting model accuracy in diverse populations. While time-varying parameters were incorporated to reflect adaptive behaviors, some parameters were assumed constant, which may not fully capture the dynamics of real-world epidemics. The use of an exponential distribution for transitions between epidemiological states simplifies the model but may not accurately represent actual waiting time distributions. Additionally, the model does not account for demographic variations, such as age/group structure and varying interaction patterns, which could provide deeper insights into disease dynamics and intervention effects. The findings from the WKU game, while valuable, may not be fully generalizable to other settings with different population dynamics and intervention strategies. Further experiments in diverse environments are needed to validate and refine the model, which is why our long-term plans would include extending the platform to support a wide range of field experiments to study transmissible diseases.

Lastly, using smartphones’ Bluetooth radio to detect proximal contacts between individuals in physical space can introduce missing data and inaccuracies. This is due to the inherent noise in the Bluetooth signal and technical issues such as varying hardware capabilities and incomplete adherence to the Bluetooth specs from smartphone makers, particularly on Android. We are currently working together with the developers of the Herald project to address these issues, and upcoming versions of the app will likely feature more accurate and robust contact detection.

### Implication for Public Health

This study underscores the importance of integrating human behavioral dynamics and NPIs into epidemiological models to improve public health responses during disease outbreaks. The experiment conducted at WKU demonstrated how real-time behavioral changes, such as adherence to social distancing, isolation and quarantine measures, can significantly alter the trajectory of an epidemic. By incorporating these adaptive behaviors into our model, we were able to provide more accurate predictions of disease transmission and identify the most effective intervention strategies. This approach emphasizes the importance of public health policies that are both responsive and adaptive, taking into account the dynamic nature of human behavior in response to changing outbreak conditions.

Also, the platform’s ability to generate comprehensive, multimodal datasets offers a valuable tool for public health planning and pandemic preparedness (for similar outbreak data generator, see [22]). The data-driven insights from this study can inform the design of targeted interventions that consider not only the biological aspects of disease spread but also the complex social and behavioral factors that influence public compliance and the effectiveness of NPIs. As public health officials navigate the challenges of controlling infectious diseases, the findings from this research provide a robust framework for developing strategies that are both evidence-based and adaptable to the changing dynamics of human behavior and pathogen transmission.

## Data Availability

All data produced are available online at https://zenodo.org/records/10674401

https://github.com/colabobio/wku-game-epi-modeling

## Declarations

### Ethics approval and consent to participate

The UMass Chan Medical School IRB determined that the research described in this manuscript is non-human research as defined by U.S. Department of Health and Human Services and Food and Drug Administration regulations (UMass Chan IRB ID: STUDY00000039); therefore there was no need for informed consent from participants of the experimental epidemic game at WKU. The protocol for the experimental game was approved by WKU Ethics Committee.

### Availability of data and source code

The analysis scripts, written in the R statistical language, are available together with usage documentation at GitHub this repository: https://github.com/colabobio/wku-game-epi-modeling, and the data from the WKU game at this Zenodo repository: https://zenodo.org/records/10674401.

## Acknowledgements

We thank the WKU students and faculty for supporting this research through their involvement in the WKU game, especially the team of WKU students who participated in the design of the activity, promoted it on campus, managed participants and provide them with information, distributed prizes, and assisted in organizing, collecting and classifying survey questionnaires (Xiangyuan Yu, Siyu Chen, He Bingze, Sixun Hou, Jinfan Jiang, Yiming Luo, HongYuan Miao, Ruiying Wang, Yueran Wang, Xuemian Xiang, and Zhang Tao). We also acknowledge Zhang Zirui for providing administrative support at WKU, Adam Fowler, creator and developer of the Herald Proximity project, Prof. Laurent Hébert-Dufresne from the Vermont Complex Systems Center at the University of Vermont for critical reading of the manuscript, and the Operation Outbreak team at the Broad Institute of Harvard and MIT, including Prof. Pardis Sabeti and Dr. Todd Brown (Operation Outbreak co-creators together with AC) and Kian Sani (Operation Outbreak product and program manager), for their support, and Fathom Information Design for the design and development of Operation Outbreak visualization tools.

## Authors’ Contributions

SSM, WM: conceptualization, data analysis, results, figures, manuscript writing; TI: figures, manuscript editing; YD, MK: user interface and graphic design; AG, HH: software development; AW, DK: planning, manuscript revision; SAN, JGY: conceptualization; AC: project lead, conceptualization, planning, manuscript writing, and funding acquisition.

---

**Figure A1:**
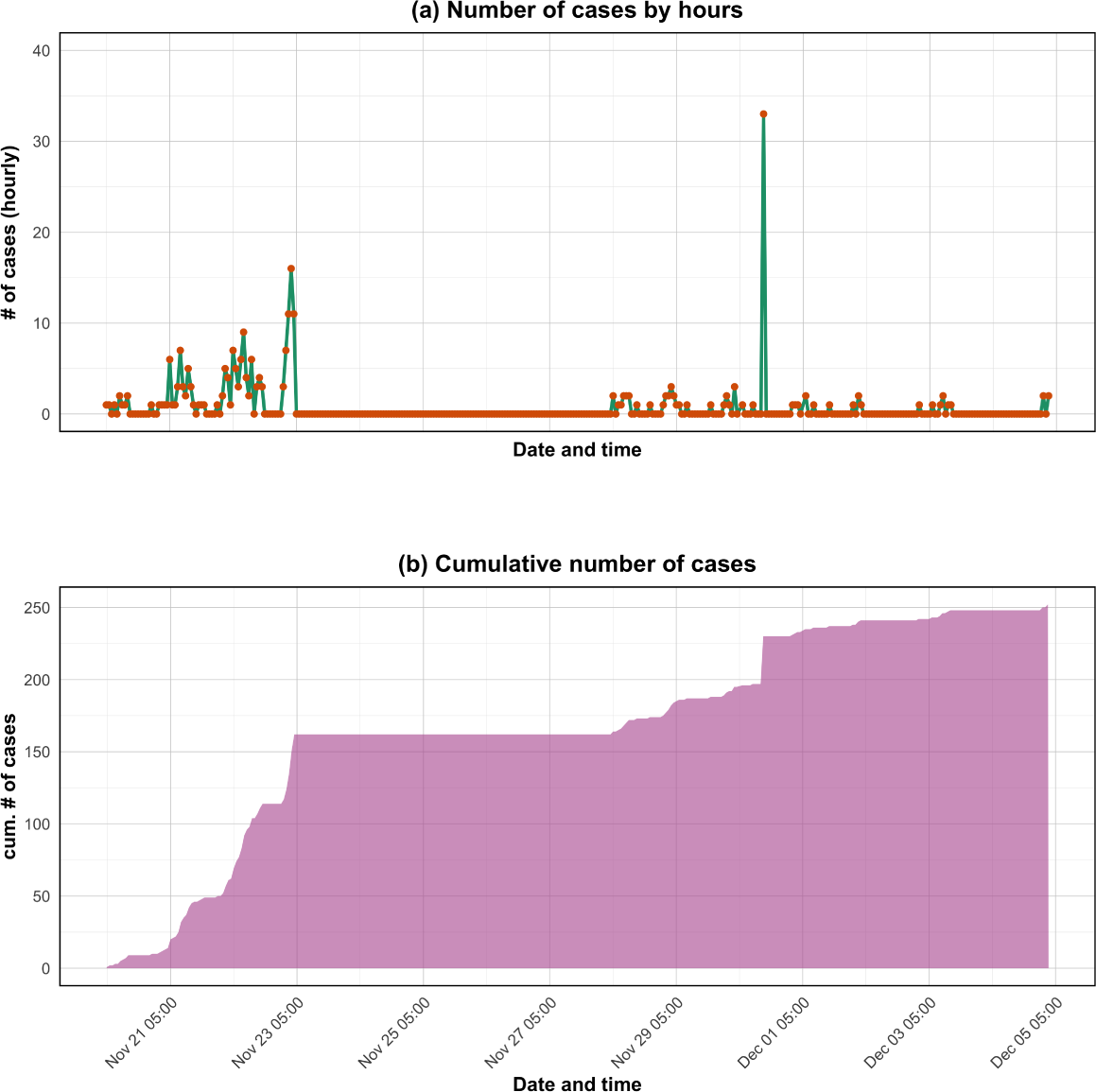
Time series plots depicting the progression of disease cases during the experiment period. The red line represents the hourly incidence data, providing a detailed view of the outbreak’s temporal dynamics. The purple line shows the cumulative number of cases, illustrating the overall growth of the outbreak over time. Specifically, panel (a) displays the hourly number of cases, capturing the shortterm fluctuations in disease transmission, while panel (b) presents the daily number of cases, offering a broader perspective on the outbreak’s progression. The four-day gap without cases corresponds to a weekend and two consecutive holidays, during which participants had limited interactions, reducing potential transmissions.

## Appendix

### A1 Time-series of the ground-truth data

The raw data generated from the WKU game (see Fig. A1) provided a comprehensive overview of the outbreak dynamics and participants’ responses. This dataset included detailed contact traces for 794 participating students over a period of 14 days, capturing crucial experiment events such as infections, re-infections, individual quarantining, and disease outcomes, including recoveries and fatalities. The sheer volume of data, totaling over 2 million individual entries, offered a granular understanding of the simulated epidemic’s progression and the effectiveness of intervention strategies.

### A2 Participants’ demographics, perceptions about quarantine, and adoption of daily protective behaviors

We administered a survey where students were able to enter their gender, major, year, mobile operating system, and other demography questions. This survey was planned to be conducted before the start of the WKU game, but due to a technical issue, this was not possible, and was distributed to participating students only several weeks after the end of the game. Because of this, only 145 students responded to the survey, which is still useful to estimate the breakdown of participation among gender, year, etc. (Fig. A2). We also provided another anonymous survey to all enrolled participants that they could optionally answer before the start of the game. This survey included three questions with an answer on a scale from 1 (strongly disagree) to 5 (strongly agree): “Public health officials should have the power to order people into quarantine during COVID-19 outbreaks”, “If someone is given a quarantine order by a public health official, they should follow it no matter what else is going on in their life at work or home”, and “If I go into quarantine, my family, friends, and community will be protected from getting COVID-19.” These questions were adapted from a telephone survey to measure public perceptions of quarantine following the H1N1 pandemic in 2009 [50]. Even though these responses were not incorporated in the current analysis either, they are part of the dataset for future analyses. Furthermore, our study aimed to capture individuals’ perceptions and actions undertaken by students based on the infection situation, offering insights into behavioral patterns and the efficacy of NPIs in mitigating disease transmission (Fig. A3). The daily behaviors of participants in the WKU game (using points to buy and wear a virtual mask or use antiviral to mitigate symptoms of disease, and choosing to quarantine/isolate for the day) were recorded by the app, and quantify the level of engagement of participants with the app as well as their concern with the possibility or consequences of infection. None of these data were used in the current analysis, as the time-varying transmission rate in our epidemiological model accounting for human behavioral responses was fitted to the case count data and did not use any of the explicit behavioral information collected with the app.

### A3 Superspreader epidemiology of the transmission tree in the WKU game

We followed the analyses described by Taube et al. in their 2022 paper [47], where they compiled the OutbreakTrees database containing 382 published and standardized transmission trees from 16 directly transmitted diseases. For each disease, they calculated several statistics, including the dispersion parameter k and mean R of the offspring distribution (the number of infections caused by each infected individual) and the proportion of cases considered superspreaders. Their analyses showed that intermediate dispersion parameters contribute most to superspreading, and provided preliminary support for the prediction that superspreaders tend to generate more superspreaders. We applied these analyses on the largest connected component of the transmission tree from the WKU game (see Fig. A4), and we were able to reproduce all their main results from published transmission trees generated by biological pathogens: (1) significant decrease in R between the first and second halves of transmission trees with 20 or more nodes and 2 or more generations of spread, (2) intermediate dispersion parameters giving rise to the highest proportion of cases causing superspreading events, such as COVID-19 (*k* = 0.14), SARS (*k* = 0.06), and MERS (*k* = 0.24), with the virtual pathogen (modeled in the app after SARS-CoV-22) yield *k* = 0.35, and (3) and the observed ratio of superspreader-superspreader exceeding what would be expected by chance in 12 of 18 trees, with COVID-19 trees showing ratios higher than 8 (the tree for the WKU game had a ratio of 14 when including all notes, and 19 when only including non-terminal nodes). These results, even though are outside the main modeling approach we use in this paper, are very important by providing strong evidence that that transmission dynamics in the WKU game is representative of biologically-caused epidemic processes, and in particular close to those of SARS-like viruses.

**Figure A2:**
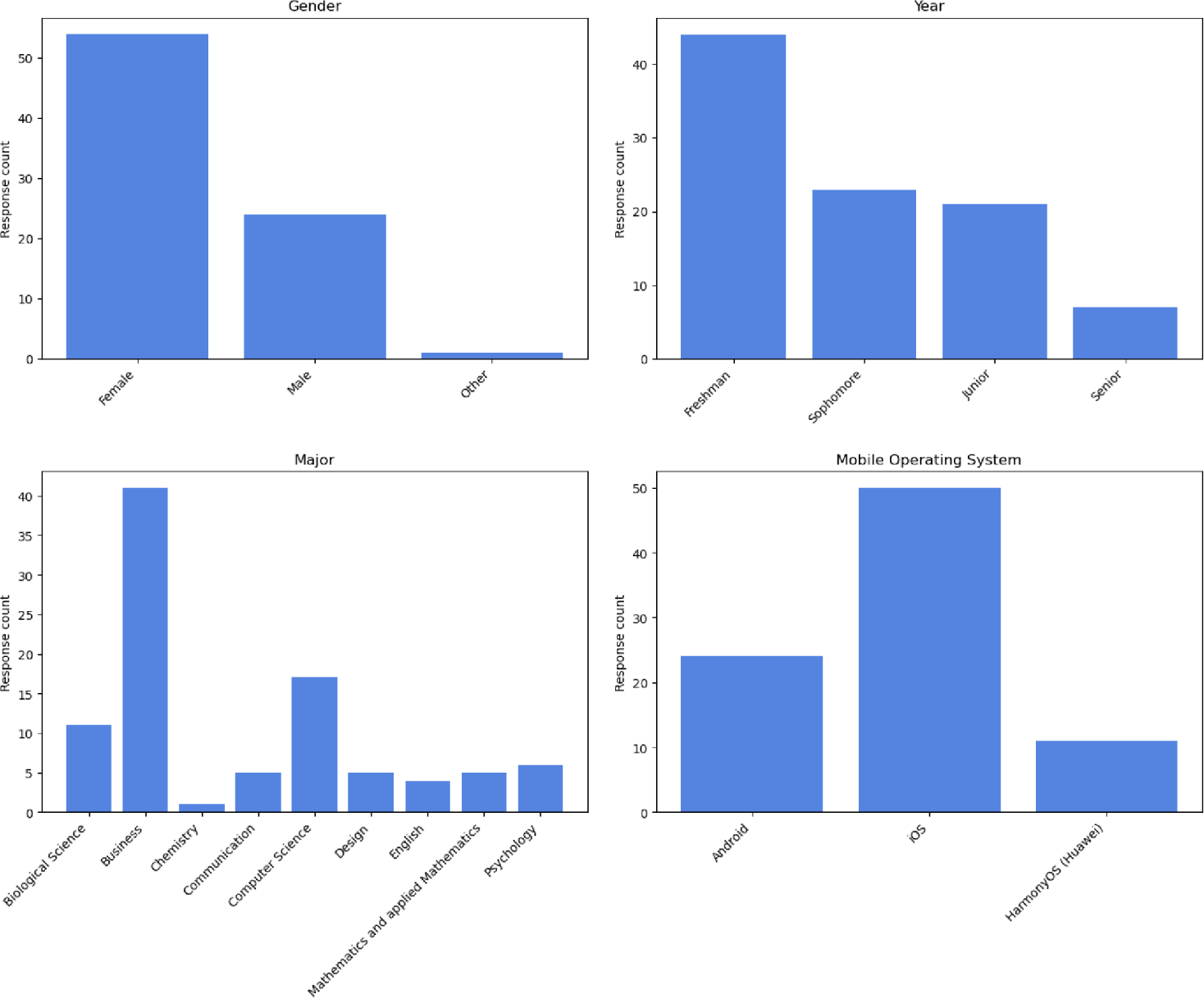
Charts representing demographics of some of the students who participated in the WKU game, collected through an anonymous survey distributed after the end of the game. These charts depict gender (female, male, other), year (freshman, sophomore, junior, senior), major (Biological Science, Business, Chemistry, Communication, Computer Science, Design, English, Mathematics and applied Mathematics, Psychology), and mobile operating system (Android, iOS, HarmonyOS/Huawei).

**Figure A3:**
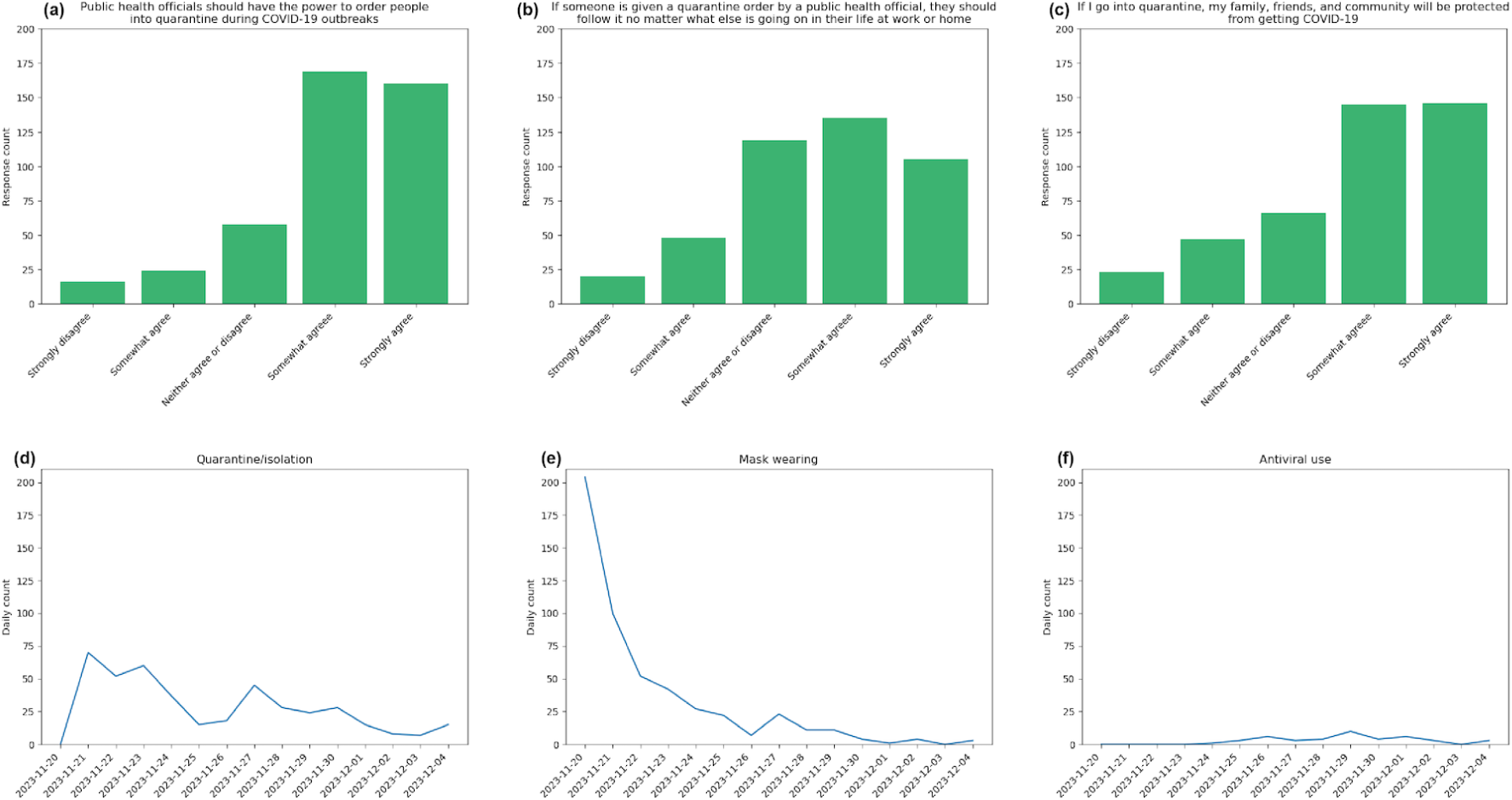
Charts depicting the perceptions of participating students regarding quarantining (top row). The height of the bars correspond to the number of students who selected each response in a scale from 1 to 5 to the three questions in the survey. The line plots show the number of students who chose to quarantine/isolate, wear a mask, and take an antiviral medication in the app at each day of the WKU game.

### A4 Model of disease transmission and progression in the app

Current epidemic games using the epidemic app are driven by a customizable model of disease transmission and progression that the app uses to calculate the probabilities of participants to move between epidemiological (e.g.: susceptible, infectious) and health (e.g.: asymptomatic, severely sick) states. A diagram of this model is depicted in Fig. A5). This model provides formulas for calculating the mean total duration of infection (broken down into exposed, infectious, and symptomatic states) and how this duration can be adjusted using rations between the different duration periods to control the pace of a experiment based on real-world infection data. Given a set of mean duration parameters, the app uses exponential distributions to randomly assign duration values during the experiment to each participant. To determine probability of infection between susceptible and infectious individuals, the model incorporates factors like estimated contact rate and duration of contact, the contact detection accuracy from the Bluetooth library, and internal time-step such that everything is normalized to that time-step for consistency. By working out the formulas in the ground-truth transmission model in the app, it is possible to link its parameters with those in a classical classical SEIR (Susceptible-Exposed-Infectious-Removed) model, such as the transmission rate, infection period, and exposure period. This enable us to run simulated scenarios in advance for each experimental game, and tweak the parameters in the ground-truth model so that the game takes place within the constrains of the experimental constrains (maximum duration, anticipated number of participants, etc.) An online reference is available detailing all the formulas used in the model [84].

**Figure A4:**
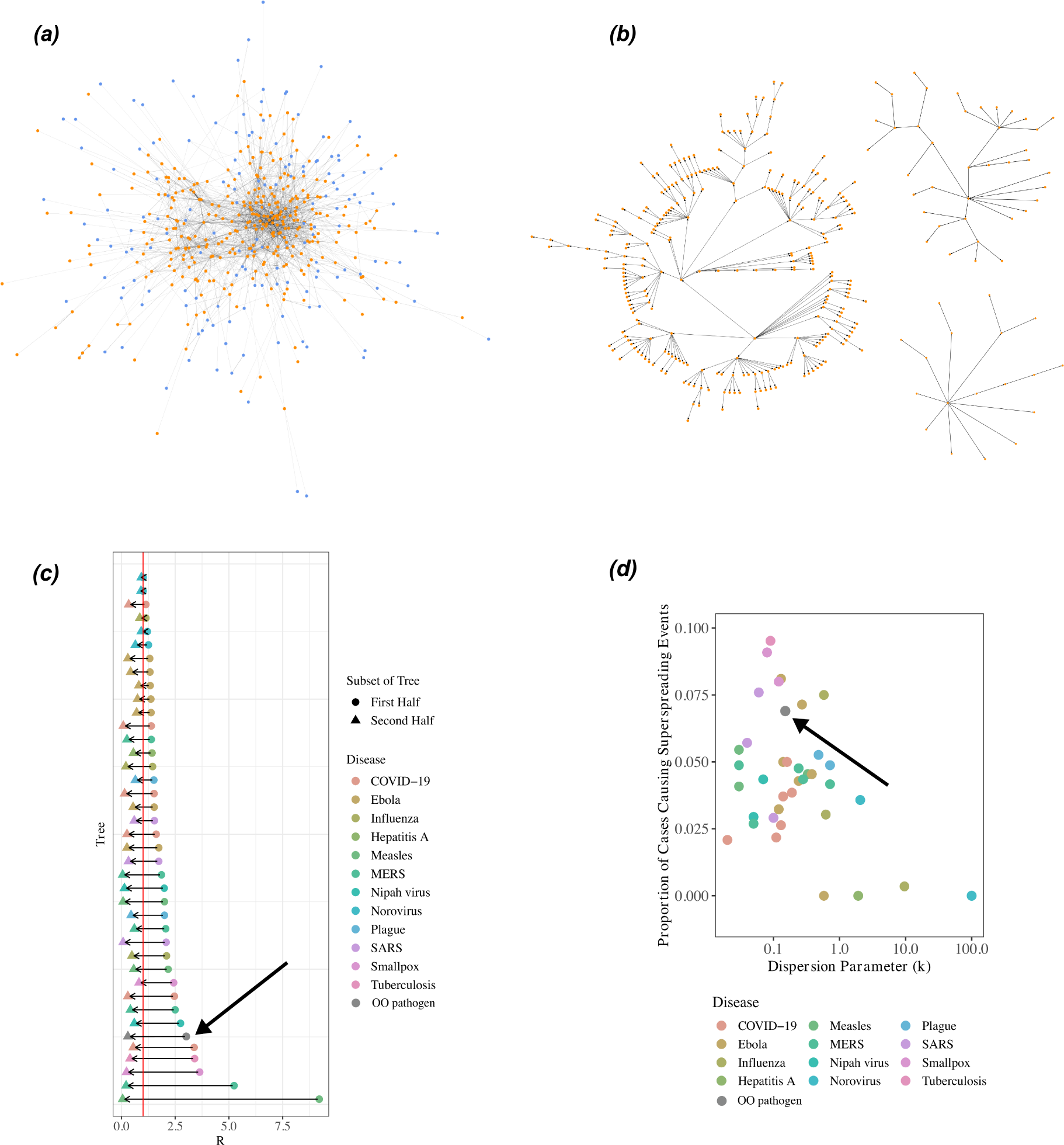
Network and superspreader epidemiology analysis of the data: contact network aggregated over the two weeks of the WKU game and colored by final infection status (blue for participants who were never infected, orange for those infected at some point during the game) (a), three largest connected transmission trees connecting infectors with infectees (b), decrease in R by disease, R was below 1 in the second half of all trees (red line denotes R = 1) (c), and proportion of cases causing super spreading events as function of dispersion parameters, as predicted by theory and measured from the data (d). Panels (c) and (d) were adapted from [47].

**Figure A5:**
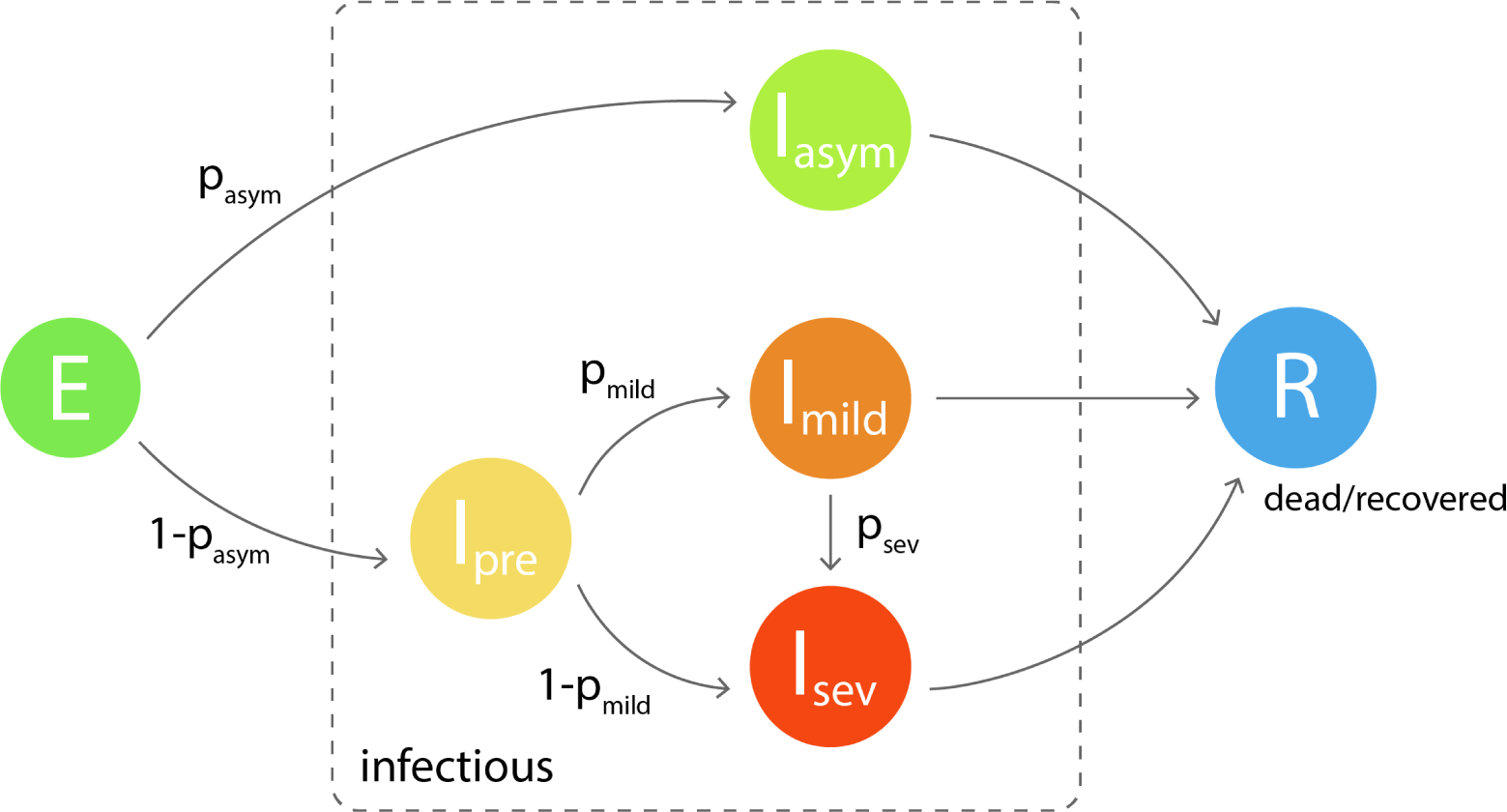
Model of disease progression in the app. “Infected” participants start in the exposed (E) state, where they are not infectious, and from there they move on to the infectious state. The infectious state contains a sequence of possible substates depending on the symptomatology of the participants; first, the participants can become fully asymptomatic but infectious (*I_asym_*) with a probability *p_asym_*. If this does not happen, with probability 1 − *p_asym_* the participants will turn into symptomatic, going through a pre-symptomatic state (*I_pre_*), which can evolve into symptomatic mild (*I_mild_*) or symptomatic severe (*I_sev_*) with probability *p_mild_* and 1 − *p_mild_*, respectively. A participant in the symptomatic mild state can become severe with probability *p_sev_*. From all these states, *I_asym_*, *I_mild_*, and *I_sev_*, the individual finally becomes removed, by either “recovering” or “dying”.

### A5 Time-2varying effective reproduction number estimation by renewal equation

The transmissibility of the pathogen in the WKU game can be quantified by calculating the instantaneous (effective) reproduction number, *R*(*t*), which represents the expected number of secondary cases generated by a single infectious individual at time *t*. In this study, we estimated *R*(*t*) from the time series data using the serial interval (SI) approach, as described by Wallinga and Teunis (2004) [71]. The SI in epidemiology refers to the time interval between successive cases in a chain of transmission [72]. With known distributions for the SI, it is possible to simulate the sequence of infections, determining the number of secondary infections caused by one primary infection based on the reproduction number. Conversely, if both the SI distribution and case time series are known, the reproduction number can be reconstructed retrospectively.

This approach to estimating *R*(*t*) has been extended in several studies [73, 74, 75, 76, 77, 78, 81] and has been applied to analyze the transmission dynamics of various infectious diseases [79, 80]. For the WKU game, we adopted the same framework to estimate the time-varying *R*(*t*) by applying the renewal equation, expressed as Eq. (A1):

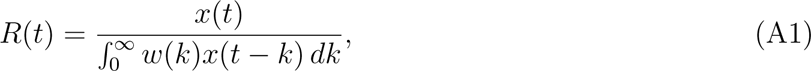

where *x*(*t*) represents the incidence rate at time *t*, and the convolution term ^f^ *^∞^ w*(*k*)*x*(*t* − *k*) *dk* measures the total infectiousness at time *t*. The term *w*(*k*) refers to the distribution of the SI, which defines the period over which an individual remains infectious. This methodology allows for a detailed and dynamic estimation of the pathogen’s transmissibility during the experiment.

Using this method, we adopted the SI estimate for COVID-19 from China as a proxy for the SI of the simulated outbreak [46]. Following approaches similar to those in [79, 80, 82, 81]. The estimated reproduction number, *R*(*t*), for the duration of the simulation, was computed, and we provided 95% confidence intervals (CI) based on Gamma priors [75, 77]. These estimates are crucial for understanding the transmission dynamics and for validating the effectiveness of non-pharmaceutical interventions (NPIs) implemented during the experiment.

The *R_eff_* is illustrated in Fig. 3, with the gray points indicating the calculated values and the error bars representing the 95% confidence intervals. A smoothed black line captures the overall trend, providing insights into the changing transmission potential over time. The red dashed line at *R_eff_* = 1 marks a critical threshold: values above this indicate the infection is spreading, while values below suggest the outbreak is under control. To estimate *R_eff_*, we utilized serial interval data from COVID-19 cases in China, scaled appropriately for the two-week duration of the experimental game [46]. This analysis offers a comprehensive understanding of the infection dynamics, identifying key transmission peaks and assessing the effectiveness of interventions in controlling the outbreak.

### A6 Mathematical formulation of the model

Following the above description, the proposed model partitions the total population *N* (*t*) at any time *t* into nine distinct sub-populations: non-quarantine susceptible (*S_nq_*(*t*)), quarantine susceptible (*S_q_*(*t*)), non-isolated exposed (*E_ni_*(*t*)), isolated exposed (*E_i_*(*t*)), non-isolated asymptomatically infected (*A_ni_*(*t*)), isolated asymptomatically infected (*A_i_*(*t*)), non-isolated symptomatically infected (*I_ni_*(*t*)), isolated symptomatically infected (*I_i_*(*t*)), recovered (*R*(*t*)), and deceased (*D*(*t*)) individuals. The model tracks the movement of individuals between these compartments using a system of non-linear ordinary differential equations (ODEs). So that:

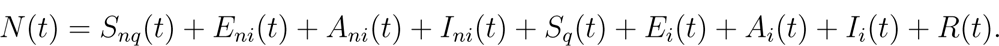

In the model, the rate at which individuals move between compartments is determined by various parameters, including the infection rate *λ*(*t*), transition rates between different states (*σ*, *γ*, *τ*), and quarantine/isolation dynamics (*ψ_ni_*(*t*), *ψ_i_*(*t*)). The model also includes parameters for recovery (*θ*), immunity waning (*ω*), and disease-induced mortality (*δ*). Individuals may leave the susceptible compartment (*S_nq_*) to become exposed (*E_ni_*) due to infection, or they may transition into or out of quarantine (*S_q_*) based on the rates *ψ_ni_*(*t*) and *ψ_i_*(*t*). Once exposed, individuals may progress to either asymptomatic (*A_ni_*) or symptomatic infection (*I_ni_*), with similar transitions for their isolated counterparts (*A_i_*, *I_i_*). Recovered individuals can lose immunity and re-enter the susceptible class. The model further accounts for simulated “deaths” due to the disease, captured by the compartment *D*(*t*).

The proposed model is depicted in Fig. 4; the state variables and model parameters (Table A1) fulfill the following system of non-linear ordinary differential equations (ODEs):

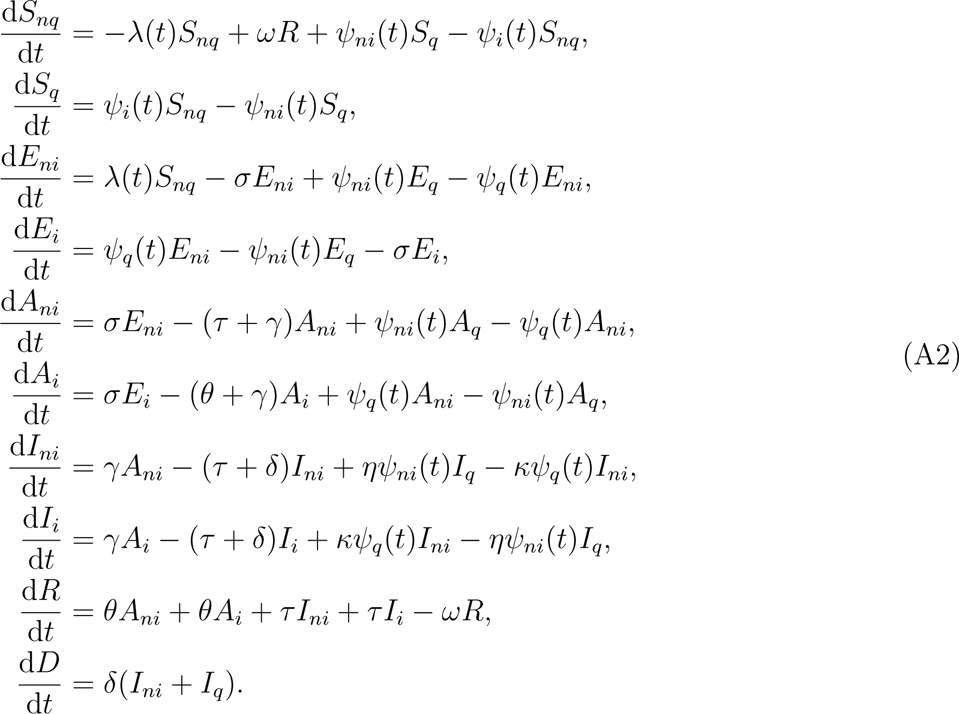

From model (A2), the incidence function or force of infection is given by the following equation:

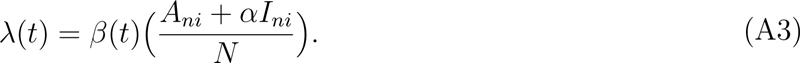

It is worth mentioning that the parameter 0 *< α <* 1 represents the adjustment factor for reduced infectiousness in infectious individuals, indicating that *A_ni_* is more prone to transmitting the disease at a higher rate than *I_ni_*. This is due to the silent nature of asymptomatic cases and the reduced activity of severely infected individuals, who are less mobile within the community compared to asymptomatic cases [57, 58]. We set the state variable *D*(*t*) to measure the number of diseasedeceased individuals to keep track of disease-related “deaths” (for calibration and quantification of model parameters).

### Model assumptions

The primary epidemiological assumptions underlying the formulation of the system (1) are as follows:

- **Heterogeneous mixing:** The population is heterogeneously mixed, reflecting varying contact patterns influenced by individual behavior, risk perception, and adherence to non-pharmaceutical interventions (NPIs). This assumption captures the complexity of real-world interactions.
- **Exponential waiting time:** Individuals transition between epidemiological states according to an exponential distribution, implying constant transition probabilities, simplifying the model while maintaining analytical tractability.
- **Continuous transitions:** Transitions between compartments occur continuously over time, accurately representing the dynamic progression of the epidemic.
- **Constant population size:** The total population remains constant throughout the experiment, focusing exclusively on disease dynamics without accounting for participants leaving the simulation for causes unrelated to their outcome within the experiment.
- **Time-varying parameters:** Key parameters such as transmission and contact rates are modeled as time-varying, reflecting adaptive behavioral responses to the evolving epidemic.
- **Behavioral response incorporation:** Human behavioral responses, including compliance with NPIs and changes in contact patterns, are integrated as dynamic variables, informed by real-time data and prior studies [55, 56].
- **Data-driven calibration:** Model calibration is based on empirical data from the WKU game, ensuring alignment with observed transmission dynamics and enhancing predictive accuracy [19, 21].
- **Feedback mechanisms:** The model includes feedback loops where behavioral changes influence, and are influenced by, the disease dynamics, capturing the reciprocal relationship between human behavior and pathogen transmission.

These assumptions offer a robust framework for comprehending and modeling disease dynamics in a controlled setting—the WKU setting in this instance—and also help us understand intervention strategies and behavioral reactions, which offers crucial insights into containing the outbreaks.

#### A7 Analysis of simplified model

The simplified version of the model (depicted in Fig. A6 and formulated in (1)), which maintains the same incidence function as in the full model (A2), while using flexible transmission rate.The simplified version of the model (1) was modified with two additional classes: (1) “*P* “ mimicking the public perception of risk regarding the number of infection (severe or critical) cases; and (2) “*C*” representing the number of cumulative cases. Note that it is also convenient to define the cumulative rate of change of disease infection as *C_sub−model_* = *σE*+*γA.* Since quarantined and isolated individuals do not contribute to disease spread, the force of infection remains unaffected. Thus, *λ*(*t*) is given by 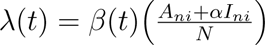, where *α* accounts for the reduced infectiousness.

##### A7.1 Basic reproduction number

Here, we computed the basic reproduction number (R_0_) of the simplified model at diseasefree equilibrium (and by considering constant transmission rate *β*_0_) by adopting the next-generation matrix (NGM) technique as demonstrated in [60]. The R_0_ represents the number of secondary cases that a typical primary case would cause during the infectious period in a wholly susceptible population [60, 61, 62, 63].

Through direct calculation of the NGM, we obtained R_0_ as follows:

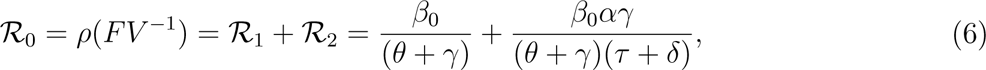

where the parameter *ρ* represents the spectral radius of the next-generation matrices of the Jacobin matrices.

Therefore, the basic reproduction number, R_0_, can be interpreted by decomposing it into three components as follows: *R*_1_, representing new infections from asymptomatically infected contacts, and *R*_2_, representing new infections from symptomatically infected contacts. This breakdown allows for a more nuanced understanding of how different types of infections contribute to the overall transmission dynamics of the outbreak. Understanding R_0_ and its components is crucial for assessing the potential impact of an infectious disease outbreak. A high R_0_ indicates that each infected individual is, on average, infecting more than one other person, leading to exponential growth in cases. This highlights the importance of interventions targeting both asymptomatic and symptomatic cases to reduce R_0_ and control the spread of the disease effectively.

The result of Theorem A7.1 below follows from Theorem 2 of [60], and reference to the local stability of the DFE of model (1), the result of Theorem 3.1 below follows.

**Theorem A7.1.** The disease-free equilibrium of model (1) *is locally-asymptotically stable whenever* R_0_ ≤ 1.

The epidemiological importance of the given theorem indicates that a small infection of the virus would not result in a substantial epidemic if the basic reproduction number (*R*_0_) is lower than one. Attaining a *R*_0_ value of less than 1 is efficacious, although not obligatory, in averting significant epidemics. When the value of *R*_0_ is below one, the sickness naturally diminishes, but it continues to exist when *R*_0_ is more than 1. This highlights the need for sophisticated intervention techniques to successfully manage the disease, as demonstrated in earlier studies like [61].

**Figure A6:**
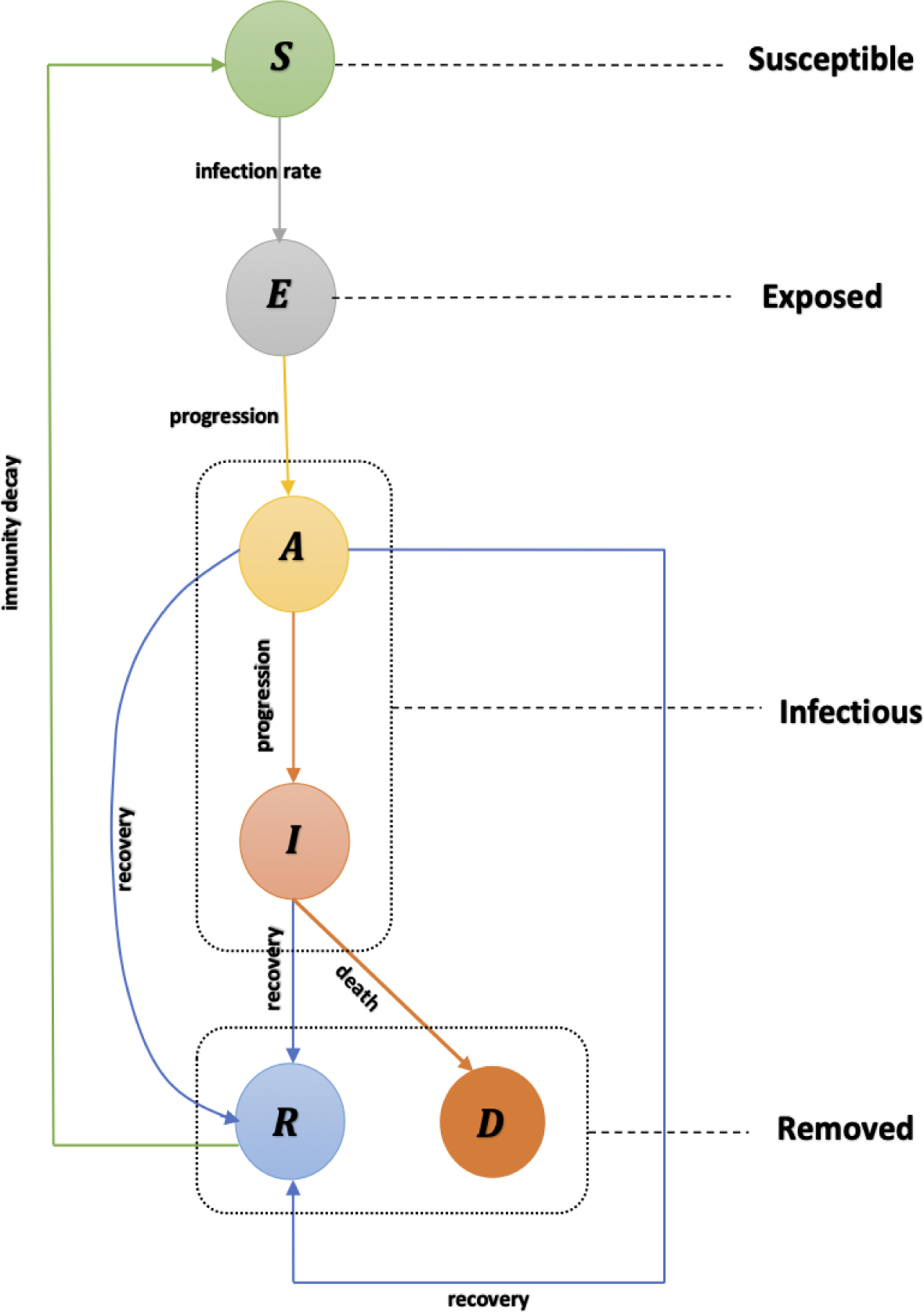
A simplified diagrammatic representation of the sub-model, derived from the full model (1), focusing on the essential dynamics of pathogen transmission. Solid arrows represent transitions between compartments, with *per capita* flow rates indicated alongside each arrow. The sub-model includes key compartments: susceptible (*S*), exposed (*E*), asymptomatic (*A*), symptomatic (*I*), and recovered (*R*) individuals. This streamlined version did not consider quarantine/isolation as separate compartment but rather as a function in the transmission rate to reduce the complexity when analyzing behavioral responses, providing a clearer view of the primary transmission pathways. While simplified, this model effectively captures the core mechanisms of disease spread, making it suitable for foundational analysis and rapid simulations aimed at understanding the impact of basic intervention measures.

#### A7.2 Model fitting and parameter estimation

In this study, we employed a compartmental SEIR model to fit the observed data on disease transmission dynamics during the WKU game. Our fitting process utilized the simulation inference framework, leveraging Pearson’s Chi-square and the least squares method to align our model with the data. This comprehensive approach involved conducting 10,000 random simulations to explore the parameter space thoroughly, ensuring an accurate representation of the outbreak dynamics [64, 65, 66]. The experimental data, which captured pathogen transmission among 794 participants over two weeks, provided a robust basis for parameter estimation. Initial conditions and demographic parameters were derived from the experiment context. Key epidemiological parameters, such as transmission and reinfection rates, were estimated to reflect the observed dynamics accurately. To estimate the parameters of the model, we conducted a fitting process where parameters such as *β* (transmission rate), *α* (modification parameter for the increase/decrease of infectiousness of infectious individuals), and *ω* (rate of reinfection) were sampled uniformly within specified ranges. This process was iterated 1,000 times, and for each iteration, the model was solved using the lsoda function from the deSolve package in R. The predicted number of cases (*C_sub−model_*) was compared to the data observed in the experiment, and the fitting accuracy was evaluated using the Pearson chisquared statistic. The iteration yielding the lowest chi-squared value was considered the best fit. Additionally, we computed the 95% confidence interval for the fitted model to assess the uncertainty in the predictions. The results of our model fitting, illustrated in Fig. A7, show a close alignment between observed cumulative incidence and model predictions, validating the model’s accuracy. The estimated parameters, detailed in Table A2, highlight the impact of human behavior and intervention strategies on disease spread.

To quantify the goodness of fit, we calculated the R-squared statistic, which represents the proportion of variance in the observed data explained by the model. The R-squared value is computed using the formula:

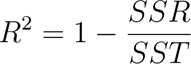

where SSR is the sum of squares of residuals (the difference between observed and predicted values) and SST is the total sum of squares (the difference between observed values and their mean). The R-squared value provides an indication of how well the model captures the variability in the observed data, with values closer to 1 indicating a better fit. In our fitting process, we achieved an R-squared value of 0.87, suggesting that the model accurately represents the observed disease dynamics. In the subsequent sub-section, sensitivity analyses were further assessed, which underscored the importance of timely interventions and compliance with preventive measures. These findings demonstrate the utility of our platform in generating valuable epidemiological insights, emphasizing the role of behavioral factors in controlling outbreaks and informing public health strategies.

**Figure A7:**
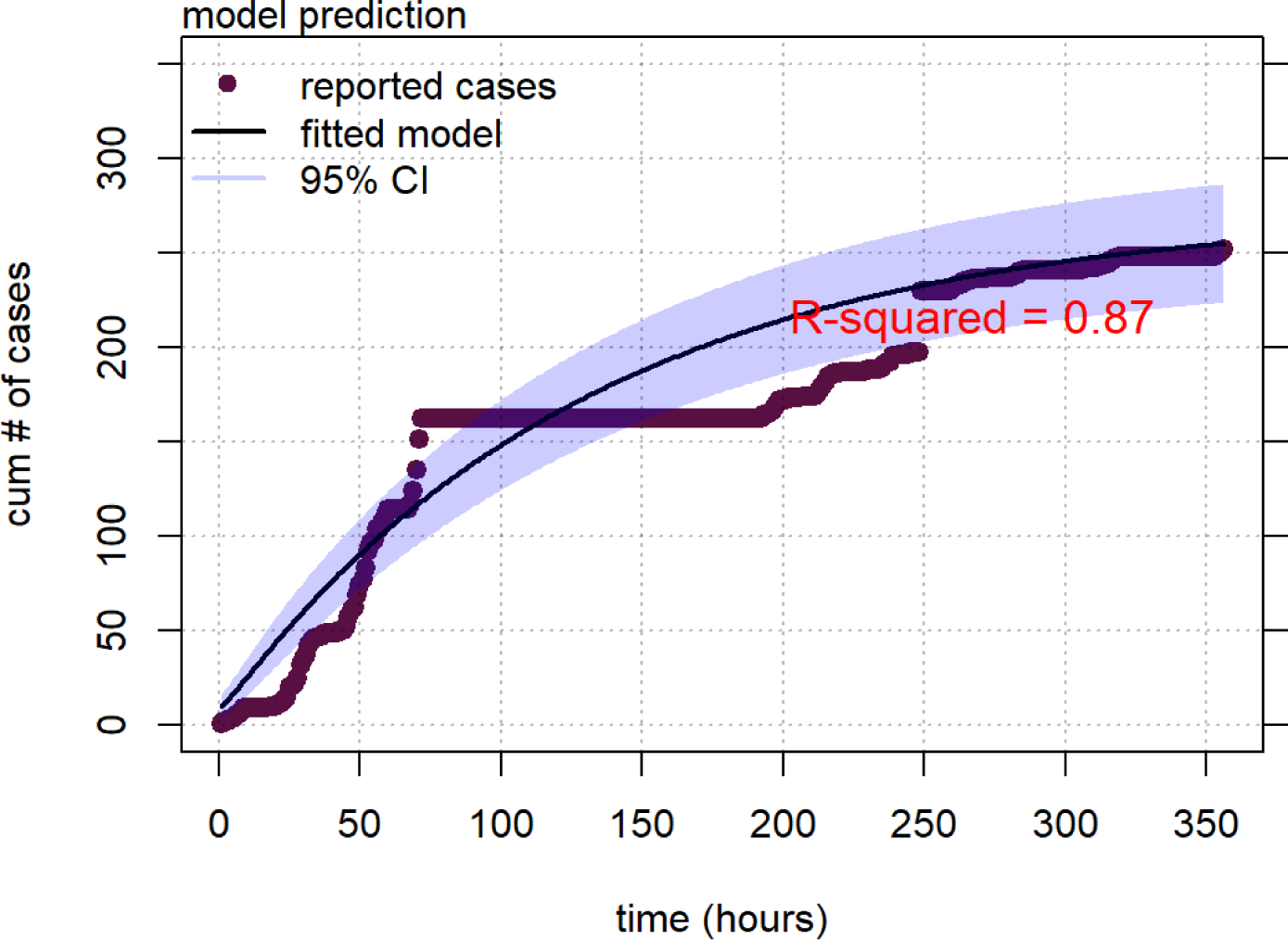
Model (6) fitting for transmission dynamics in the WKU game. The gray dotted points represent the observed data, while the red curve depicts the model’s predictions based on the fitted epidemic parameters. The light-blue shaded area indicates the 95% credible interval, reflecting the model’s uncertainty range. This analysis was conducted using R Statistical Software version 4.2.1. The specific parameters employed in the simulation are detailed in Table A2

#### A7.3 Further Numerical Simulations

Furthermore, we conducted further numerical simulations to gain valuable insights into how varying key epidemiological parameters, such as transmission rate (*β*), recovery rate (*θ*), and progression rate (*γ*), influence the overall dynamics of disease spread. By testing 50% increases and decreases in these parameters, we can assess their impact on infection patterns.

Figures A8(a) and A8(b) illustrate the effect of varying *β* on asymptomatic (*A_n_*) and symptomatic (*I_n_*) infections. In Figure 1(a), a 50% increase in *β* leads to a higher proportion of asymptomatic cases over time, while a 50% decrease reduces the number of asymptomatic individuals. This highlights the importance of minimizing transmission rates to control outbreaks. Similarly, Figure A8(b) shows that a 50% increase in *β* results in a substantial rise in symptomatic cases, whereas a 50% reduction significantly decreases symptomatic infections. These results emphasize the critical role of interventions aimed at reducing transmission (e.g., social distancing, mask-wearing) in mitigating both asymptomatic and symptomatic spread.

Figures A8(c) and A8(d) explore the influence of recovery (*θ*) and progression rates (*γ*) on disease dynamics. In Figure A8(c), increasing *θ* by 50% leads to a faster reduction in asymptomatic infections, while a 50% decrease prolongs the infectious period, maintaining higher levels of transmission. This underscores the importance of enhancing recovery rates through early detection and treatment. In Figure A8(d), a 50% increase in *γ* accelerates the transition from asymptomatic to symptomatic cases, increasing the overall burden of symptomatic infections. Conversely, a 50% reduction in *γ* delays the progression, reducing the number of symptomatic cases over time. This highlights the significance of interventions that slow disease progression to alleviate the outbreak’s severity. These simulations demonstrate the profound effect that modifying *β*, *θ*, and *γ* has on controlling disease transmission, underscoring the importance of targeted interventions in outbreak management.

**Figure A8:**
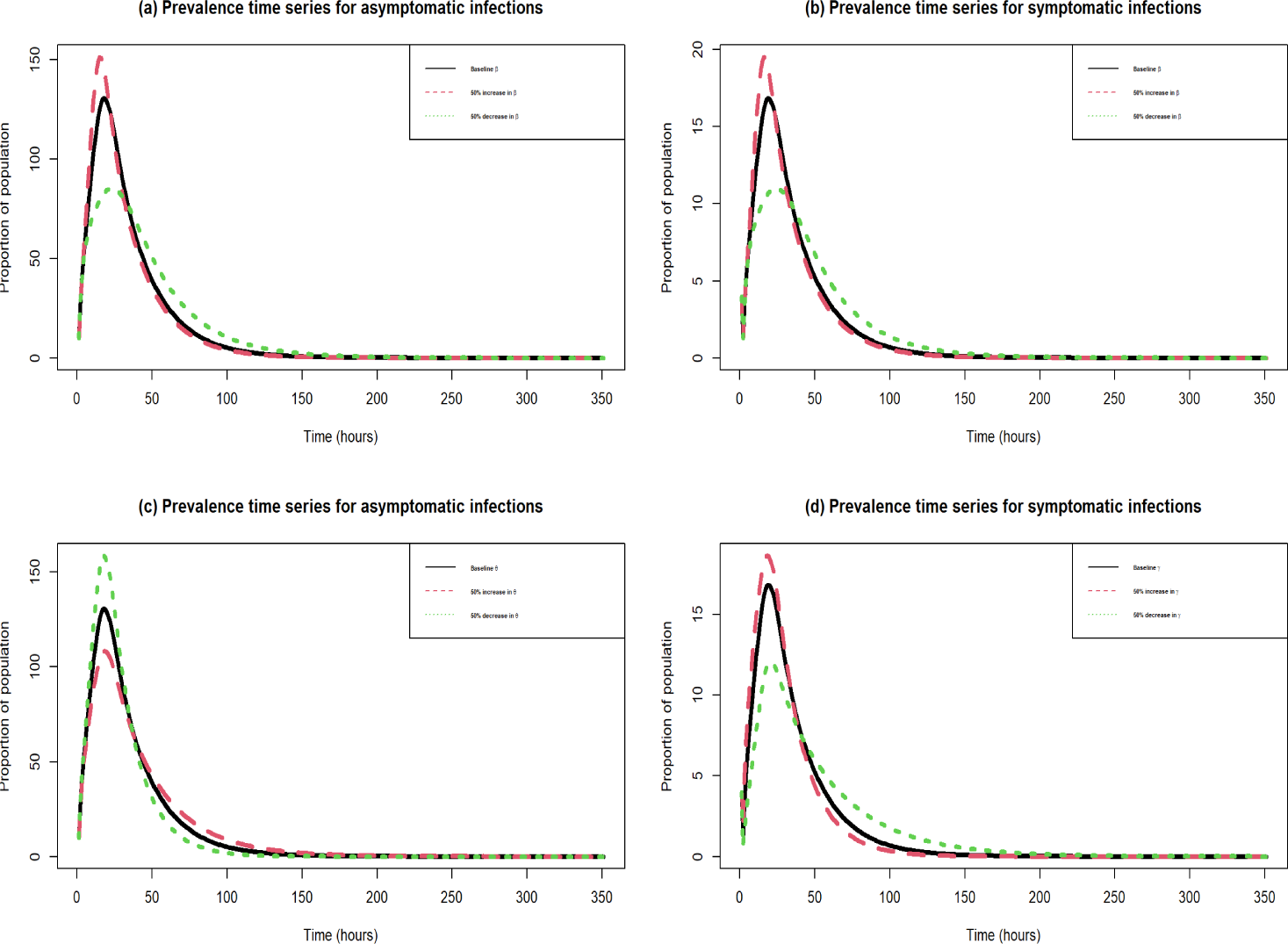
Solution curves of disease dynamics for varying transmission rates *β*, recovery rates *θ*, and progression rates *γ*. (a) Prevalence time series for asymptomatic infections (*A*) with varying transmission rates (*β*). The plot shows the effect of baseline, 50% increase, and 50% decrease in *β* on the proportion of asymptomatic individuals over time. (b) Prevalence time series for symptomatic infections (*I*) with varying transmission rates (*β*). The plot illustrates how baseline, 50% increase, and 50% decrease in *β* impact the proportion of symptomatic individuals over time. (c) Prevalence time series for asymptomatic infections (*A*) with varying recovery rates (*θ*). The plot shows the effect of baseline, 50% increase, and 50% decrease in *θ* on the proportion of asymptomatic individuals in the population over time. (d) Prevalence time series for symptomatic infections (*I*) with varying progression rates from asymptomatic to symptomatic infections (*γ*). The plot demonstrates the effect of baseline, 50% increase, and 50% decrease in *γ* on the proportion of symptomatic individuals over time. The legends in each subplot clearly indicate the different variations in *β*, *θ*, and *γ*, highlighting the influence of these key parameters on the infection dynamics within the population.

#### A8 Summary table of initial state values and parameters estimation results of the model **(1)**

**Table A1:** Epidemiological description of the state variables and parameters of model (1).

| Variable | Description |
| --- | --- |
| $S_n$ | Non-quarantine susceptible |
| $S_q$ | Quarantine susceptible |
| $E_n$ | Non-isolated exposed |
| $E_i$ | Isolated exposed |
| $A_n$ | Non-isolated asymptomatic (mild or no symptoms) |
| $A_i$ | Isolated asymptomatic |
| $I_n$ | Non-isolated symptomatic (moderate to severe symptoms) |
| $I_i$ | Isolated symptomatic |
| $R$ | Recovered |
| $P$ | Risk perception |
| Parameter | Description |
| $\beta_0$ | Probability of transmission per contact |
| $\alpha$ | Modification parameters for decreased infectiousness |
| $\sigma$ | Transition rate of exposed individuals to asymptotically infected |
| $\gamma$ | Progression rate from asymptomatic to symptoms stages |
| $\psi_q$ | Rate of movement from non-quarantine/non-isolation to quarantine/isolation |
| $\psi_n$ | Rate of movement from quarantine/isolation to non-quarantine/non-isolation |
| $\eta$ | Rate of reduced transition from isolation to non-isolation symptomatic individuals |
| $\kappa$ | Rate of reduced transition from non-isolation to isolation for symptomatic individuals |
| $\theta$ | Recovery rates of symptomatically infected individuals |
| $\tau$ | Recovery rates of symptomatically infected individuals |
| $\omega$ | Immunity waning rate |
| $\delta$ | Disease-induced death rate |
| $\epsilon$ | Rate of intervention action strength |
| $\zeta$ | Response intensity (measures the intensity of response) |

**Table A2:** Summary of parameter values for model (1).

| Parameter | Default value (range) | Units | Sources |
| --- | --- | --- | --- |
| $\beta_0$ | 0.8366158 (0.25 – 1.5) | per time | fitted |
| $\alpha$ | 0.489124 (0.4 – 0.6) | per time | assumed |
| $\sigma$ | 0.126 (0.01 – 0.25) | per time | simulated |
| $\gamma$ | 0.129 (0.05 – 0.2) | per time | simulated |
| $\psi_i$ | 0.06 (0.042 – 0.09) | per time | [65, 68] |
| $\psi_{ni}$ | 0.06 (0.042 – 0.09) | per time | [65, 68] |
| $\eta$ | 0.6333 (0.001 – 0.95) | per time | fitted |
| $\kappa$ | 0.084 (0.05 – 0.1) | per time | simulated |
| $\theta$ | 0.18 (0.1 – 0.4) | per time | [68, 65] |
| $\tau$ | 0.017 (0.05 – 0.2) | per time | [68, 65] |
| $\omega$ | 0.2411152 (0.1 – 0.75) | per time | assumed |
| $\delta$ | 0.983 (0.1 – 0.95) | per time | assumed |
| $m$ | 1/1.5 (0.1 – 0.92) | per time | assumed |
| $\epsilon$ | 0.1 (0.05 – 0.275) | per time | simulated |
| $\zeta$ | (0.1 – 0.5) | per time | simulated |

